# Resolution limit-free community detection reveals unique patterns of resting-state network connectivity in posttraumatic stress disorder: A PGC-ENIGMA PTSD Consortium investigation

**DOI:** 10.1101/2021.06.24.21259102

**Authors:** Marisa C. Ross, Josh M. Cisler, Saskia B.J. Koch, Miranda Olff, Dick J. Veltman, Laura Nawijn, Jessie L. Frijling, Mirjam van Zuiden, Xi Zhu, Yuval Neria, Benjamin Suarez-Jimenez, Tor Wager, Courtney Haswell, Michael D. De Bellis, Emily Clarke Rubright, Jennifer S. Stevens, Sanne J.H. van Rooij, Negar Fani, Tanja Jovanovic, Kerry J. Ressler, Judith K. Daniels, Henrik Walter, Antje Manthey, Anika Sierk, Pavel Riha, Ivan Rektor, Richard Davidson, Jack B. Nitschke, Dan Grupe, Christine Larson, Terri deRoon-Cassini, Jacklynn Fitzgerald, Ashley Huggins, Carissa Weis, Ruth Lanius, Maria Densmore, Lauren A. M. Lebois, Milissa L. Kaufman, Justin T. Baker, Thomas Straube, Paula Neumeister, David Hofmann, Amit Etkin, Adi Maron-Katz, Anthony King, Israel Liberzon, Michael Angstadt, Ryan Herringa, Xin Wang, Tian Chen, Andrew Cotton, Brian O’Leary, Hong Xie, Seth Disner, Nicholas Davenport, Wissam El Hage, Yann Quidé, Elbert Geuze, Mitzy Kennis, Evan Gordon, Geoffery May, Steven Nelson, Meilin Jia-Richards, Steven Bruce, Ilya Veer, Lea Waller, Hannah Berg, Shmuel Lissek, Rajendra A. Morey, Scott Sponheim

## Abstract

Posttraumatic stress disorder (PTSD) is a complex psychiatric condition that has generated much attention in the neuroimaging literature. A neurocircuitry model supporting fronto-limbic dysfunction as a major player in facilitating clinical symptoms of PTSD is well-characterized; however, recent literature suggests that network-based approaches may provide additional insight into neural dysfunction in PTSD. Our analysis uses resting-state neuroimaging scans of 1063 adults from the PGC-ENIGMA PTSD Consortium to investigate a network-based model of functional connectivity in PTSD. With a novel, resolution limit-free community detection approach, 16 communities corresponding to functionally meaningful networks were detected with high quality. After group-level community detection, participants were classified into three groups (PTSD, *n*=418, trauma-exposed controls without PTSD, *n*=434, and non-trauma exposed healthy controls, *n*=211). Individual network connectivity metrics were calculated, including whole-brain, default mode network, and central executive network participation coefficient and connectivity strength. Linear mixed effects models revealed group differences in the whole-brain, default mode, and central executive network participation coefficient and connectivity strength such that individuals with PTSD demonstrated overall greater values. We also described sex differences such that males demonstrate greater whole-brain participation coefficient vs. females and females demonstrate greater default mode network connectivity strength vs. males. Our results suggest that PTSD in adults is associated with reduced specialization and enhanced inter-module communication throughout the brain network, which may contribute to inefficient information processing and poor emotional regulation. This study presents a novel use of resolution limit-free community detection in a large PTSD sample, revealing robust differences in resting-state network topology.

## 1. Introduction

The complex nature and high burden of posttraumatic stress disorder (PTSD), has merited extensive research into its neurobiological bases. Through the use of emotion- and threat-processing functional magnetic resonance imaging (fMRI) tasks, a neural model of PTSD that centers dysfunction in the fronto-amygdala circuit has emerged. This model, informed largely by rodent models of fear conditioning ^1–3^, posits that PTSD symptoms and related deficits arise from a hyperactive amygdala and hypoactive ventral medial prefrontal cortex (vmPFC) comprising a circuit, independent from other brain circuits, that facilitates behaviors such as enhanced fear expression, failures in extinction learning, attention bias to threat, and heightened arousal ^4–6^. Though this model is supported by rigorous scholarship, contemporary work has identified inconsistencies in fMRI research that suggest the fronto-amygdala circuit may only be a small part of the complete story of neural deficits in PTSD ^7–10^. More recent work proposes the involvement of several other neural circuits that show consistent deficits in PTSD, including hippocampal-vmPFC ^6,11^ and dorsal anterior cingulate cortex-anterior insula ^6^ circuits. Further, a recent review ^12^ of functional connectivity studies in PTSD failed to find convincing support for the fronto-limbic model but rather revealed patterns of functional connectivity in PTSD that reflect distinct large-scale functional networks differences. As such, network-based investigations may complement traditional univariate and bivariate approaches and provide valuable insights into altered neural function in trauma-exposed clinical samples.

Network-based investigations of mental health disorders, including PTSD, conceptualize the brain as a dynamic series of connections with distinct functional sub-networks that work in tandem to produce coordinated brain activity. While several functional components have been studied in the context of psychiatric disorders, three are described in the “Triple Network Model” of psychopathology ^13^. The three networks in this model show coupled activity during tasks and resting state ^13,14^ and include the salience network (SN), central executive network (CEN), and default mode network (DMN). The SN, anchored in the anterior insular cortex and dorsal anterior cingulate cortex ^15^, is important for mapping internally and externally salient events and switching between cognitive systems accordingly ^13,16^. Alterations in connectivity of the SN may contribute to hyperarousal symptoms in PTSD. Primary regions of the CEN include the dorsal lateral prefrontal cortex and posterior parietal cortex ^15^. Because the CEN is thought to facilitate control of spatial attention ^17,18^ and emotional regulation ^19,20^, cognitive impairments in PTSD patients including attentional bias to threat and deficits in emotional regulation ^21,22^ may be driven by changes in the connectivity of this network. Finally, the DMN is anchored in the posterior cingulate cortex and mPFC and facilitates self-referential mental activity and regulation of emotional states ^13,23^. Altered connectivity of the DMN, which also contains the hippocampus ^23^, may affect contextual processing ^11^, which may contribute to re-experiencing symptoms and deficits in fear extinction in PTSD.

Network-based methods allows testing of hypotheses about brain function of individual brain regions (“nodes” of the graph) and connections between them (“edges”), individual functional components (“communities”), and whole-brain functional organization in relation to psychiatric symptoms. Various graph theory concepts can be applied to characterize nodes and edges within functional brain networks, including node-level participation coefficient (PC) and connectivity strength. The PC describes the extent to which a node’s edges are distributed throughout other communities in the graph ^24^ and is a measure of inter-community connectivity ^25^ similar to modularity. When averaged across a network or community, the value of the PC reflects a measure of integration of communication, with a value approaching zero representing a graph with edges contained almost entirely within their own communities and a value approaching one representing near uniformity of connections across all communities ^24^. In PTSD, one hypothesis may be that low PC contributes to enhanced specialization of fear-processing networks, while high PC may indicate disorganized or inefficient processing of emotional information, especially within emotion regulation centers such as the CEN, DMN, and SN. Connectivity strength is calculated based on the correlation in functional activity between nodes and represents a statistical history of co-activation between two regions ^26^. This edge-level metric can be averaged across the graph or within a community to represent the strength of communication between brain regions across a graph or within a community. In PTSD, one hypothesis regarding connectivity strength may be that it is either too weak - contributing to breakdowns in information processing - or too strong, contributing to inefficient communication across and hyper-specialization of emotion-processing communities.

Before graph-based measures can be calculated, a method to detect distinct communities (also referred to as networks or modules) within the graph must be applied. While several methods exist for determining community structure in graphs, few have been applied to PTSD samples and many have substantial limitations. In functional brain networks, a pressing concern is the resolution limit of many community detection methods ^27,28^, including the popular Newman’s Modularity ^29^. When using common community detection algorithms with resolution limits, researchers must select a resolution parameter that defines the minimum size of communities that can be resolved within the graph. This process determines the number of distinct communities that are detected in the graph; therefore, allowing for smaller communities results in the detection of more communities and requiring larger communities results in detecting fewer communities. Consequently, experimenter bias is imposed upon an otherwise data-driven technique through the selection of this resolution parameter.

Conversely, community detection approaches that lack a resolution limit provide promising new avenues for detecting an anatomically-meaningful community structure in functional brain networks that has less potential for bias than data-driven techniques for selecting a resolution parameter. Importantly, calculation of graph metrics such as the PC depends on the number and size of the resolved modules in the network ^30^; therefore, a resolution limit-free approach can also yield node-level graph metrics with reduced bias. Though resolution limit-free community detection has yet to be used in a PTSD sample, an application to patients with schizophrenia ^31^ suggests that these methods can be applied to resting-state brain networks to reveal clinically relevant novel differences in network topology.

The following analysis utilizes the PGC-ENIGMA PTSD Consortium dataset and a novel resolution limit-free community detection approach to identify a network-based model of PTSD. Whole-brain and community-specific average PC are examined to characterize differences in network integration and connectivity strength. Our goal was to characterize communication strength across the brain and within the CEN, DMN, and SN, which were of interest given their critical role in emotion regulation ^13,19,20,23^. This analysis investigates differences in PC and connectivity strength in relation to PTSD diagnosis, PTSD symptom severity, index trauma type, and a binary designation of sex. Index trauma type was of interest because incidence of PTSD varies greatly depending on the type of trauma experienced, with interpersonal violence (IPV) conferring the highest risk ^32^. We test the hypothesis that this differential incidence may be partially explained by changes in network topology dependent on trauma exposure type. Additionally, because PTSD is more prevalent in females compared to males ^33–35^ and recent evidence suggests males and females demonstrate divergent connectivity patterns in resting-state networks and small-worldness ^36,37^, binary sex was also included as a predictor of interest in all analyses. Ultimately, we aimed to characterize a new, data-driven model of functional connectivity differences in PTSD that may serve as a springboard for future research on improving outcomes for patients.

## 2. Methods

### 2.1 Samples

Clinical and resting-state functional neuroimaging data from 1233 individuals were collected from 25 cohorts assessed in 20 different laboratories across six countries. Please see **Tables S1 and S2** for cohort-level demographic information and inclusion criteria. Participants were either non-trauma exposed controls (NTC), trauma-exposed controls without PTSD (TEC) or met diagnostic criteria for PTSD. Current PTSD was assessed using the Clinician Administered PTSD Scale for DSM-IV (CAPS-IV ^38^) in twelve cohorts, the CAPS for DSM-5 (CAPS-5 ^39^ in seven cohorts, the PTSD Checklist for DSM-IV (PCL-C ^40^) in two cohorts, the PCL for DSM-5 (PCL-5 ^41^) in two cohorts, and one cohort each used the DSM-IV symptom scale and the Posttraumatic Stress Diagnostic Scale^42^. Because assessments varied across sites, symptom severity was calculated within each cohort as a percentage of total possible score on the given assessment, resulting in a range of scores from 0-1. Individuals selected for analysis were between the ages of 18 and 55 years.

### 2.2 Image Acquisition and Preprocessing

Resting-state image acquisition parameters varied across the different cohorts. Repetition times ranged from 1.3 to 3 seconds and resting-state scan times ranged from five to eight minutes. Technical parameters such as voxel size, spatial resolution, number of slices, and slice thickness also varied across sites (**Table S3**). To minimize any potential systematic differences resulting from acquisition parameters, only sites utilizing a 3-Tesla MRI scanner and gradient echo EPI acquisition sequences were included.

Resting-state images were acquired at each site and preprocessed at a single location (Duke University). Preprocessing was completed through the ENIGMA HALFpipe workflow (https://github.com/HALFpipe/HALFpipe^43^) which relies on fMRIPrep^44^. Briefly, HALFpipe was used to complete steps motion correction, slice timing correction, susceptibility distortion correction, and spatial normalization. fMRIPrep calculates the parameters for each transformation, applying them in one step and extracting all confound signals before denoising with ICA AROMA^45^. Denoising is completed after resampling functional data to standard space in fMRI prep, defining the standard space as the MNI152NLin2009cAsym template. Denoising occurs in several steps, first with ICA AROMA, which classifies component timeseries as signal or noise, followed by spatial smoothing with AFNI’s 3dBlurInMask^46^, followed by grand mean scaling to set the mean image, removal of noise components, low-and high-pass temporal filtering, and removal of nuisance time series through residualization. Images were initially smoothed with a 6.0 mm FWHM kernel in the HALFpipe workflow, then resmoothed with an 8.0 mm FWHM kernel using AFNI’s 3dBlurToFWHM^46^ before the following analyses to improve comparability of spatial normalization across the many cohorts in this analysis.

### 2.3 Group-Level Connectivity Matrices

A 250-regions-of-interest (ROI) functional atlas from Craddock and colleagues ^47^ was applied to each participant’s resting-state timecourse. The mean timecourse of voxels within each ROI for each participant was calculated, excluding voxels within ROIs that were outside of the brain for a given individual, resulting in a 250×250 symmetrical connectivity matrix for each participant. ComBat harmonization ^48,49^ in Matlab was applied to connectivity matrices to correct for systematic scanner effects. Covariates included group (PTSD, TEC, or NTC), PTSD symptom severity, binary sex, index trauma type, and age to ensure that ComBat would not remove variance of interest for this analysis.

Harmonized matrices were concatenated along a third dimension across participants, r-to-z transformed, all diagonals were set to zero, and the mean was taken within each ROI across participants. Only ROIs that were common across all participants were included, resulting in 29 ROIs being excluded and a 221×221 group-level connectivity matrix representing functional connectivity between the same ROIs across all participants. In line with recommended sparsification procedures ^50^, percolation analysis was used to determine the optimal sparsification threshold. Percolation analysis refers to an iterative process where connections are removed from a graph until the largest community begins to break down ^51^ and is used to ensure a large, sparse graph with noisy connections removed while maintaining every node is connected to at least one other node. Following determination of the proper threshold, the connectivity matrix for the group and for each subject were independently thresholded and carried forward for community detection.

### 2.4 Resolution Limit-Free Community Detection

Large-scale network structure was determined using a contemporary approach for resolution limit-free community detection based on the probability distribution of Asymptotical Surprise (AS). AS is a weighted extension of the Surprise parameter, which represents the improbability of finding an observed graph partition by chance in a random graph ^52^. Surprise estimates a null distribution that assumes that links between nodes of a graph appear randomly, then evaluates the distance between the null distribution and the given partition to provide a measure of graph partition quality ^53^. AS is preferred to Surprise in brain networks because it reliably estimates the quality of partitioning in weighted (i.e. non-binarized) graphs ^50^. As such, AS can be used to investigate community structure of brain networks that is unbounded by a resolution limit, thereby enabling a unique characterization of network organization in PTSD.

The Partitioning Cost Optimization (PACO) algorithm ^50^ in Matlab was used to maximize AS and perform community detection. PACO has recently been applied to resting-state fMRI data in healthy adults and schizophrenia patients to circumvent the resolution limit of most community detection algorithms ^31,54^ and has outperformed other community detection methods^50^. PACO is a non-deterministic, agglomerative algorithm that uses the Jaccard Index ^55^ to agglomerate similar nodes of the graph into communities by moving single nodes between communities in the graph and merging similar nodes until AS stops improving ^50^. Because PACO does not rely on a resolution parameter to detect communities, it can identify small, neurophysiologically relevant communities within large brain networks. PACO was run 10,000 times on the thresholded, group-level connectivity matrix and the iteration with the maximum AS value was chosen as the optimal graph partition for the group. Please see **Figure 1** for a flowchart of image processing and community detection steps.

**Figure 1.**
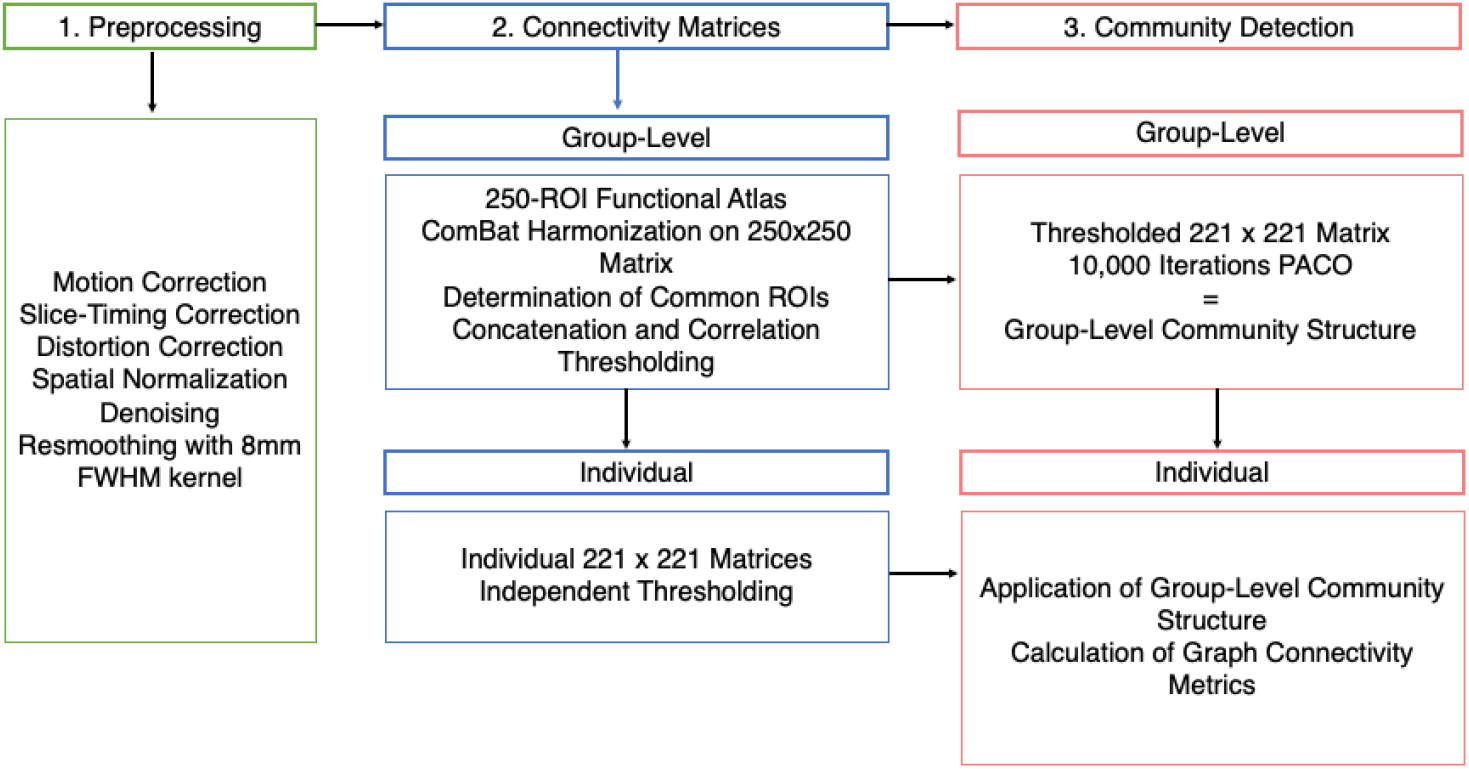
Flow-Chart of Resting-State Image Processing and Community Detection. All resting-state scans were preprocessed through the HalfPipe workflow at the Duke University site (1). Group-level correlation matrices were generated, thresholded, and harmonized after application of a 250-ROI functional atlas from Craddock et al. and determination of common ROIs. The common ROI structure was applied to individuals for independent thresholding (2). Group-level community structure was determined with the Partitioning Cost Optimization (PACO) algorithm and carried forward to individuals for calculation of graph connectivity metrics (3).

Jackknife analysis investigated whether any individual site or cohort was reducing quality of group-level community detection. Cohorts were removed from the final analysis if excluding the cohort increased the total number of common ROIs across all subjects *and* if Surprise values improved when that cohort was excluded from community detection. This process resulted in three cohorts being removed from the final analysis (**Table S4, Figure 2**).

**Figure 2.**
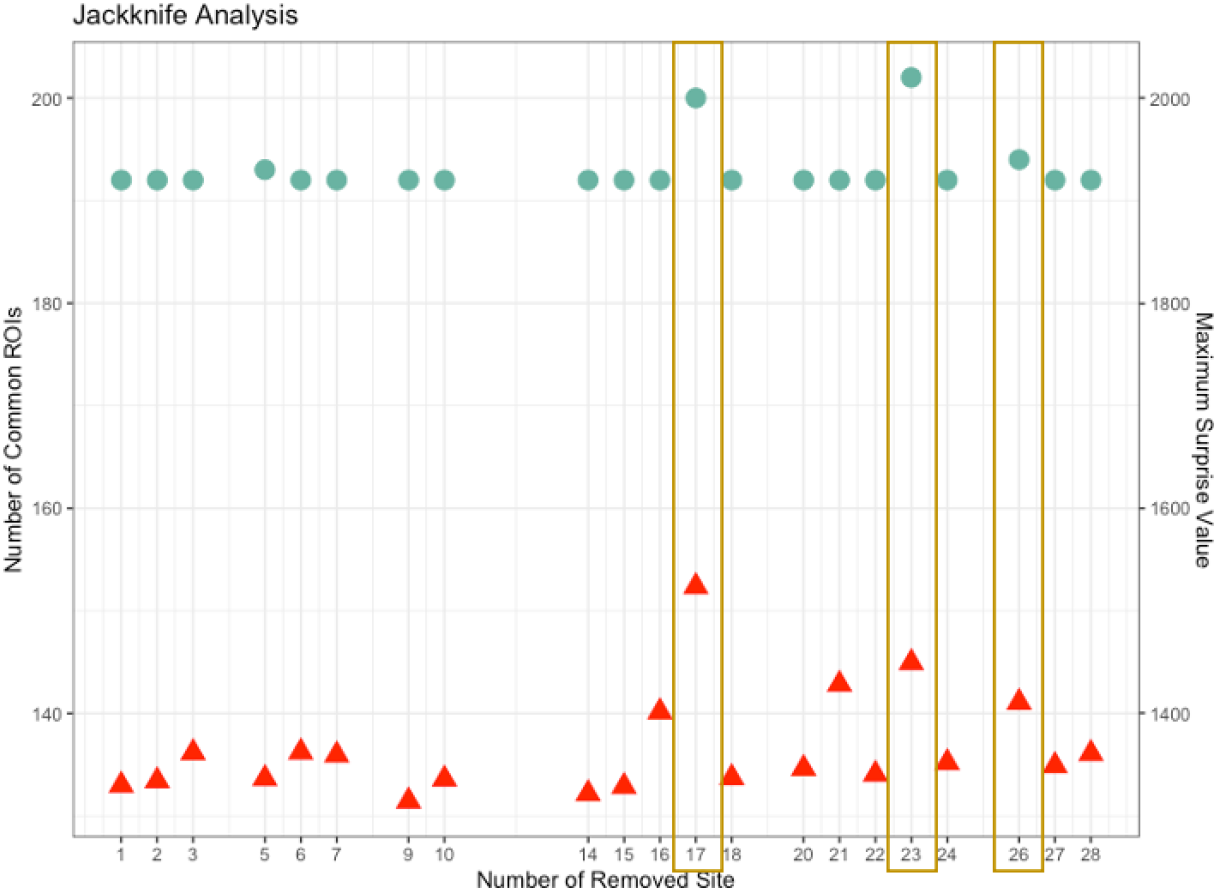
Jackknife Analysis for Quality Control. Three sites were excluded after jackknife analysis when *both* ROI coverage *and* Surprise values improved after removing each site. Red triangles represent maximum surprise value when a given cohort was removed from the group analysis; green circles represent the number of common ROIs in the group-level matrix when a given cohort was removed from the group analysis.

### 2.5 Selection of Network Communities

The optimal group-level community structure was inspected for anatomical consistency with canonical functional brain network modules ^56^. Communities consistent with the DMN and CEN according to the Yeo et al. parcellation ^56^ were the communities of interest for this analysis and therefore were carried forward for group-level analyses. While the SN was also a community of interest, PACO failed to identify a canonical SN and therefore connectivity of this network could not be investigated.

### 2.6 Calculation of Graph Connectivity Metrics

The optimal group-level community structure was next applied to each subject-level connectivity matrix to ensure comparability of network metrics across participants. The Brain Connectivity Toolbox in Matlab ^57^ was used to calculate PCs for each ROI for each subject, after which the values for each ROI were averaged to generate average whole-brain, DMN, and CEN PCs for each subject. Average whole-brain connectivity strength was calculated from each subject’s connectivity matrix by isolating the upper triangle of each subject’s symmetrical matrix to generate a vector representing connectivity strength of a given node to the rest of the brain. The mean of each vector was then taken to represent average whole-brain connectivity strength for each subject. This process was repeated within the focal communities (DMN and CEN).

### 2.7 Data Analysis

Group-level differences in whole-brain, DMN, and CEN PC and connectivity strength were determined through three sets of linear mixed effects models (LMEMs; Matlab fitlme function) testing three separate questions. All whole-brain models included continuous age as a covariate and included subject nested within scanner as a random effect to ensure that any remaining systematic variance between scanners not mitigated by ComBat was properly modeled. To assess for unique effects of DMN and CEN PC and connectivity strength above and beyond whole-brain influences, DMN and CEN models for all analyses also included a covariate for average strength and PC of the whole-brain network absent the community of interest. Holm-Bonferroni correction ^58^ was used to correct for multiple comparisons, with the whole-brain features for each model set considered a family with two separate comparisons (PC and connectivity strength) and the DMN and CEN as separate families with two comparisons. Therefore, the p-value threshold for significance was set at .025 (.05 / 2) for each family of tests.

#### 2.7.1 Group * Sex LMEM

To investigate the first aim of determining the relationship between trauma exposure and binary sex on graph connectivity metrics, group (NTC, TEC, and PTSD) was dummy-coded with PTSD set as the reference group. Dummy-coded group, binary sex, and their interaction were then entered into three separate groups of LMEMs predicting whole-brain and community-specific PC and connectivity strength, respectively.

#### 2.7.2 Symptom Severity * Sex LMEM

For the second aim of determining the relationship between PTSD symptom severity and binary sex on connectivity metrics, only participants with trauma exposure were included in another set of LMEMs. The scaled PTSD severity score described above, binary sex, and their interaction were then entered into three separate groups of LMEMs predicting whole-brain and community-specific PC, and connectivity strength, respectively.

#### 2.7.3 Symptom Severity * Index Trauma LMEM

Finally, the effect of index trauma type and symptom severity on global graph connectivity metrics was investigated with LMEMs. Index trauma type was divided into combat/military trauma, IPV exposure (i.e. sexual or physical assault, childhood and domestic abuse, etc.), and other (i.e. motor vehicle accidents, natural disasters, etc.) and dummy-coded with IPV as the reference group. Dummy-coded index trauma type, PTSD severity scaled score, and their interaction were then entered into three separate groups of LMEMs predicting whole-brain and community-specific PC and connectivity strength, respectively.

## 3. Results

### 3.1 Sample Characteristics

Subjects from three scanners (N=170) were removed from the dataset following jackknife analysis, resulting in a total subject number of 1063 (660 female; **Table 1**). The final sample included 418 subjects with PTSD (267 female; 63.8%), 434 TEC (260 female; 59.9%), and 211 NTC (133 female; 63.0%) from 21 cohorts at 16 different sites. Index trauma type was highly correlated with binary sex (*r*=-.501), with more males experiencing combat-related trauma and more females reporting interpersonal violence. Groups differed in age (*F*(2,1060)=5.456, *p*=.004) such that the NTC group was younger, on average, compared to the TEC (*p*=.005) and PTSD (*p*=.011) groups.

**Table 1:** Sample Demographics. *PTSD = posttraumatic stress disorder group; TEC = trauma-exposed control group; NTC = non-trauma exposed control group; IPV = interpersonal violence;* NTC group is younger, on average, compared to TEC and PTSD; PTSD group has higher severity than both TEC and NTC; TEC higher than NTC; *Index trauma information is missing from 191 trauma-exposed subjects

|  | PTSD | TEC | NTC | Group Difference | P Value |
| --- | --- | --- | --- | --- | --- |
| **N** | 418 | 434 | 211 |  |  |
| **N Female (%)** | 267 (63.8%) | 260 (59.9%) | 133 (63.0%) | $F(2,1060) = 0.761$ | 0.4680 |
| **Age (years)** | $32.8 \pm 8.1$ | $33.0 \pm 9.2$ | $30.7 \pm 8.7$ | $F(2,1060) = 5.456$ | 0.0044 |
| **Age range** | 18-50 | 18-55 | 18-51 |  |  |
| **PTSD Severity (range 0-1)** | $0.512 \pm 0.165$ | $0.200 \pm 0.210$ | $0.100 \pm 0.127$ | $F(2,1060) = 497.9$ | < 0.0001 |
| **Index Trauma\* (N Female)** |  |  |  |  |  |
| **Combat** | 125 (6) | 111 (19) | | $F(1,234) = 0.888$ | 0.3470 |
| **IPV** | 218 (210) | 53 (49) | | $F(1,269) = 0.242$ | 0.6230 |
| **Other** | 75 (51) | 128 (91) | | $F(1,201) = 0.585$ | 0.4450 |
| **N scanners** | 14 | 11 | 11 |  |  |

### 3.2 Group-Level Network Structure

PACO detected 16 communities from the 221×221 group-level connectivity matrix with high graph partition quality (Surprise =1799.2; **Figure 3**). The largest communities included a bilateral somatomotor network, a bilateral DMN, a bilateral visual processing network, and a bilateral CEN. Smaller communities included 1) bilateral hippocampus and parahippocampus, 2) bilateral striatum and thalamus, 3) bilateral orbital frontal gyrus, and 4) bilateral anterior insula. The communities corresponding to the DMN and CEN were *a priori* communities of interest and therefore were carried forward to group-level analyses.

**Figure 3.**
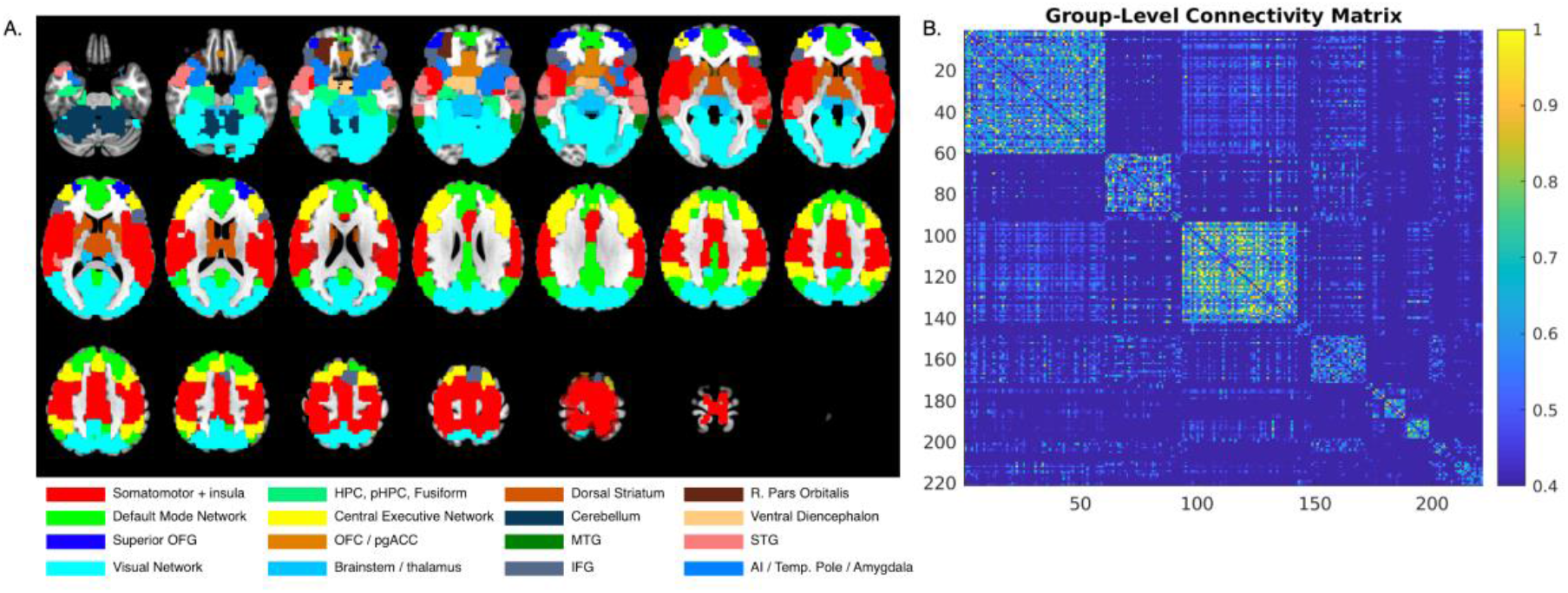
Group-Level Resting-State Network Structure. 16 distinct communities corresponding to functional networks were defined using a resolution limit-free community detection approach through the Partitioning Cost Optimization algorithm ^50^ (A). Two-dimensional group-level connectivity structure representing node-by-node connectivity patterns in the resting-state graph (B). *OFG = orbital frontal gyrus; HPC, pHPC = hippocampus / parahippocampus; OFC / pgACC = orbital frontal cortex / perigenual anterior cingulate cortex; MTG = middle temporal gyrus; IFG = inferior frontal gyrus; STG = superior temporal gyrus; AI / Temp. pole = anterior insula / temporal pole*

### 3.3 Relationship between Trauma Exposure, PTSD, and Graph Connectivity

#### 3.3.1 Average Whole-Brain PC

LMEMs testing the interactive relationship between group (NTC, TEC, and PTSD) and binary sex on the average whole-brain PC revealed group differences. A main effect of group revealed a higher average PC for the PTSD group compared to the TEC group (*t*(1056)=-3.24, *p*=.001; **Table 2, Figure 4A,B**), as well as a significant main effect of binary sex such that average PC was higher in males compared to females (*t*(1056)=2.93, *p*=.003; **Figure 4C**).

**Table 2:**
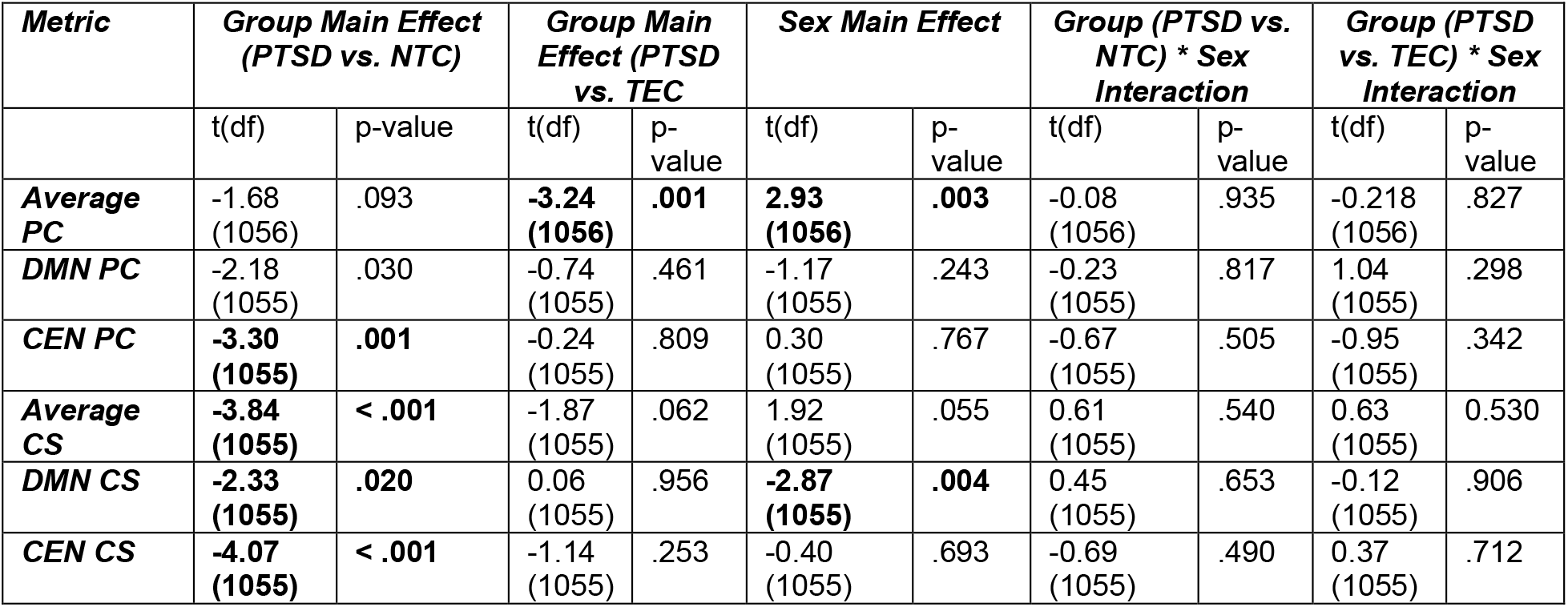
**Group-level statistics from linear mixed effects models predicting network metrics from clinical group and binary sex**. *PC = participation coefficient; CS = connectivity strength; DMN = default mode network; CEN = central executive network; PTSD = posttraumatic stress disorder clinical group; NTC = non-trauma exposed control group; TEC = trauma-exposed control group*. ***Bolded*** *values in table indicate significant differences after correction for multiple comparisons*.

| *Metric* | *Group Main Effect (PTSD vs. NTC)* |  | *Group Main Effect (PTSD vs. TEC)* |  | *Sex Main Effect* |  | *Group (PTSD vs. NTC) \* Sex Interaction* |  | *Group (PTSD vs. TEC) \* Sex Interaction* |  |
| --- | --- | --- | --- | --- | --- | --- | --- | --- | --- | --- |
|  | t(df) | p-value | t(df) | p-value | t(df) | p-value | t(df) | p-value | t(df) | p-value |
| **Average PC** | -1.68<br>(1056) | .093 | **-3.24**<br>**(1056)** | **.001** | **2.93**<br>**(1056)** | **.003** | -0.08<br>(1056) | .935 | -0.218<br>(1056) | .827 |
| **DMN PC** | -2.18<br>(1055) | .030 | -0.74<br>(1055) | .461 | -1.17<br>(1055) | .243 | -0.23<br>(1055) | .817 | 1.04<br>(1055) | .298 |
| **CEN PC** | **-3.30**<br>**(1055)** | **.001** | -0.24<br>(1055) | .809 | 0.30<br>(1055) | .767 | -0.67<br>(1055) | .505 | -0.95<br>(1055) | .342 |
| **Average CS** | **-3.84**<br>**(1055)** | **< .001** | -1.87<br>(1055) | .062 | 1.92<br>(1055) | .055 | 0.61<br>(1055) | .540 | 0.63<br>(1055) | 0.530 |
| **DMN CS** | **-2.33**<br>**(1055)** | **.020** | 0.06<br>(1055) | .956 | **-2.87**<br>**(1055)** | **.004** | 0.45<br>(1055) | .653 | -0.12<br>(1055) | .906 |
| **CEN CS** | **-4.07**<br>**(1055)** | **< .001** | -1.14<br>(1055) | .253 | -0.40<br>(1055) | .693 | -0.69<br>(1055) | .490 | 0.37<br>(1055) | .712 |

**Figure 4.**
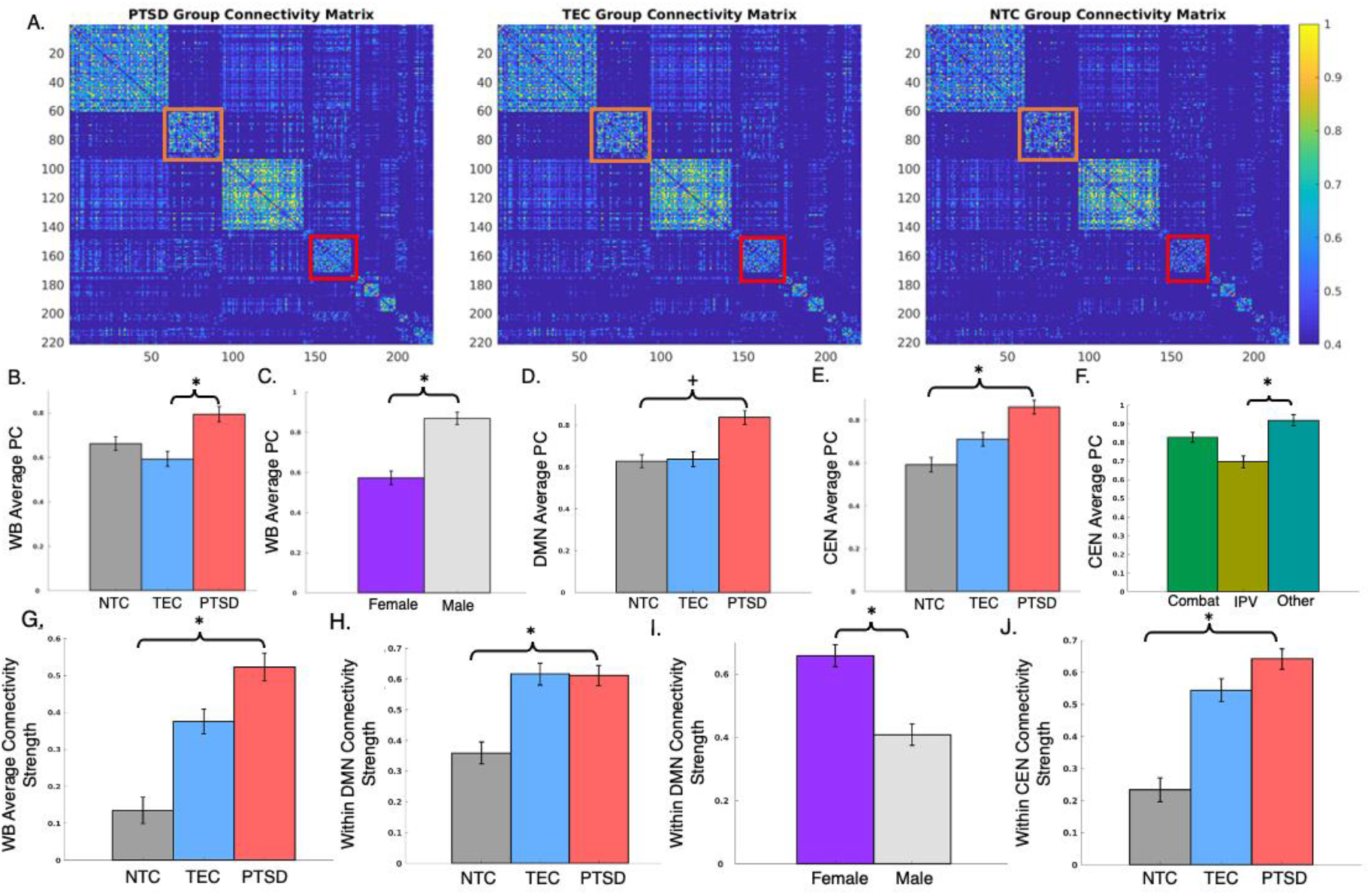
Group Differences in Resting-State Network Metrics. Individuals with PTSD showed greater whole-brain and community-level participation coefficient (PC) and connectivity strength compared to controls, which is evident in the two-dimensional connectivity matrices for each group. Orange boxes outline the DMN, red boxes outline the CEN in each group. Color bar indicates strength of correlation between nodes on scale of *r* = 0.4 to *r* = 1 (A). Whole-brain PC differed by group and binary sex with the PTSD group and males showing greater PC compared to TEC (B) and females (C), respectively. Group differences in DMN PC did not survive correction for multiple comparisons (D), while CEN PC differed significantly between the PTSD and NTC groups (E). Individuals who experienced interpersonal violence also showed reduced CEN PC compared to other types of non-combat trauma (F). Whole-brain (G), DMN (H), and CEN (J) connectivity strength was also higher in the PTSD group compared to the NTC group. DMN connectivity strength was also higher in females compared to males (I). All bar plots represent residualized group differences. *TEC = trauma exposed control group; NTC = non-trauma exposed control group; IPV = interpersonal violence*. ^*^ *p* < .02, ^+^ *p* < .05

#### 3.3.2 Average DMN PC

No significant effects for the relationship between average DMN PC and group or sex remain after correction for multiple comparisons (**Table 2, Figure 4D**).

#### 3.3.3 Average CEN PC

A main effect of group was revealed such that the PTSD group demonstrated a higher average PC of the CEN compared to the NTC (*t*(1055)=-3.53, *p*< .001) but not the TEC (*t*(1055)=-0.24, *p*=.809) groups (**Table 2, Figure 4E**).

#### 3.3.4 Average Whole-Brain Connectivity Strength

Main effects of group on connectivity strength were revealed such that the PTSD group demonstrated higher average connectivity strength across the brain network compared to the NTC group (*t*(1056) =-3.84, *p* < 0.001; **Table 2**; **Figure 4A,G**).

#### 3.3.5 Average DMN Connectivity Strength

LMEMs testing the interactive relationship between group and binary sex on DMN connectivity strength revealed a main effect of group such that individuals with PTSD demonstrated higher DMN connectivity compared to NTC (*t*(1055)=-2.33, *p*=.020; **Table 2**; **Figure 4H**). A main effect of binary sex (*t*(1055)=-2.87, *p*=.004; **Table 2**) also suggests stronger DMN connectivity in females compared to males (**Figure 4I**).

#### 3.3.6 Average CEN Connectivity Strength

LMEMs in the CEN revealed a significant main effect of group such that individuals with PTSD demonstrated higher CEN connectivity compared to NTC (*t*(1055)=-4.07, *p*< .001; **Table 2**; **Figure 4J**).

#### 3.3.7 Node-Specific Contributions to Whole-Brain Organization

Because the whole-brain measures of PC and connectivity strength represent an average, we next wanted to investigate whether the observed group difference in PC and connectivity strength was driven by individual nodes or a global network phenomenon. When the PC and connectivity strength of each node were plotted individually, all nodes showed a general trend of a greater value in the PTSD group compared to the control groups (**Figure 5**), suggesting that the observed group differences in whole-brain network metrics were not attributable to any one node in the network.

**Figure 5.**
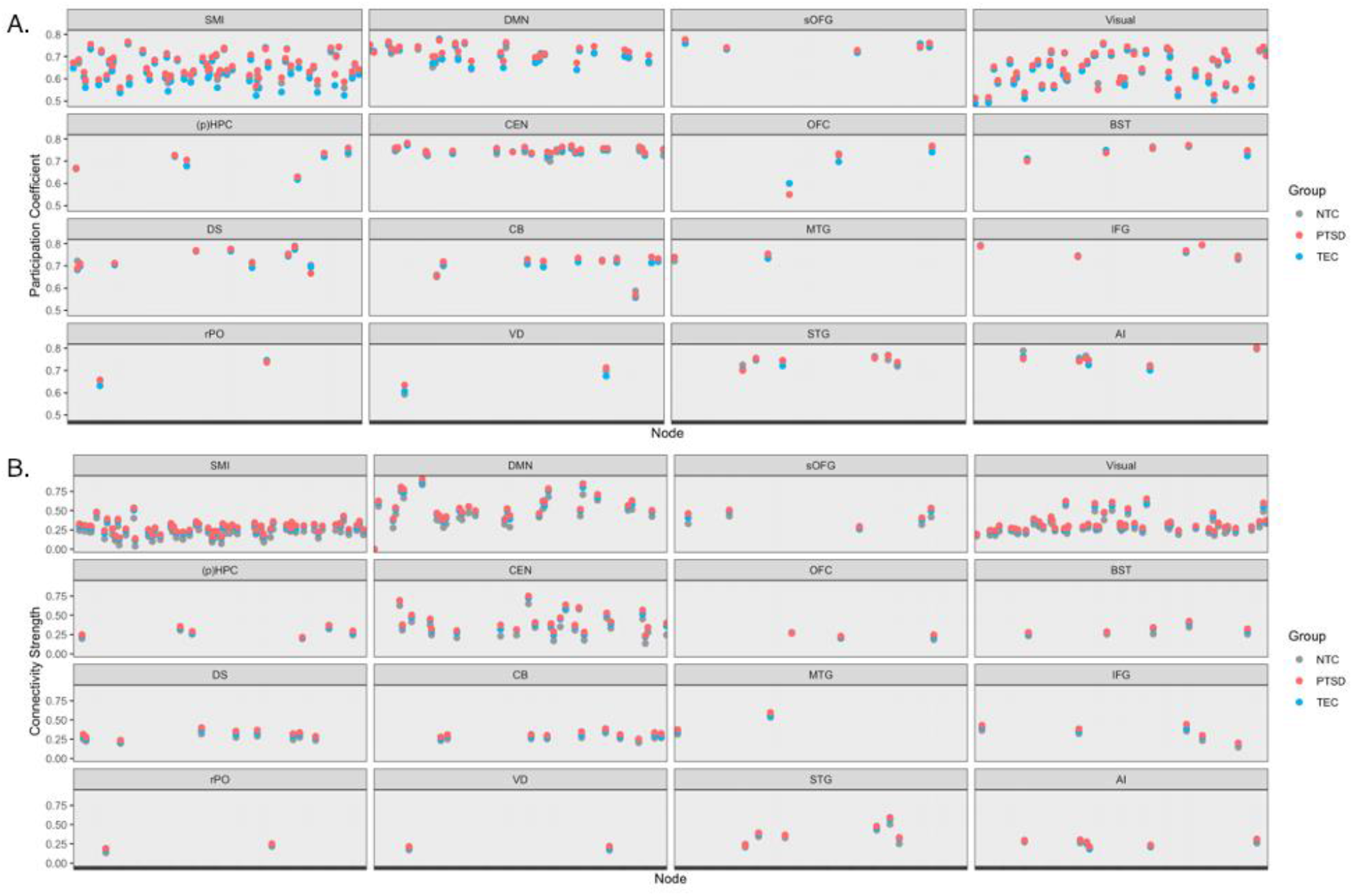
Node-Level Participation Coefficient and Connectivity Strength by Diagnostic Group. Subjects with PTSD demonstrated generally higher participation coefficient (A) and connectivity strength (B) throughout the brain graph compared to trauma-exposed controls (TEC) and in the CEN compared to non-trauma exposed (NTC) subjects, which is not driven by any one node in the graph showing particularly high participation coefficient or connectivity strength. Mean node-level measures for each group are sorted by community. Standard deviations for participation coefficient per group: NTC = 0.077, TEC = 0.084, PTSD = 0.085 (A). Standard deviations for connectivity strength per group: NTC = 0.163, TEC = 0.157, PTSD = 0.174 (B); *SMI = somatomotor and insula; DMN = default mode network; sOFG = superior orbital frontal gyrus; Visual = visual processing network; (p)HPC = hippocampus / parahippocampus / fusiform gyrus; CEN = central executive network; OFC = orbital frontal cortex; BST = brainstem / thalamus; DS = dorsal striatum; CB = cerebellum; MTG = middle temporal gyrus; IFG = inferior frontal gyrus; rPO = right pars orbitalis; VD = ventral diencephalon; STG = superior temporal gyrus; AI = anterior insula / temporal pole / amygdala*

### 3.4 Relationship between PTSD Symptom Severity and Graph Connectivity

LMEMs testing the interaction between PTSD symptom severity and binary sex on average whole-brain, DMN, and CEN PC and connectivity strength in the trauma-exposed groups failed to reveal any significant effects for the predictors of interest other than a similar main effect of binary sex such that females demonstrate stronger DMN connectivity strength compared to males (*t*(846)=-3.29, *p*=.001; **Table 3**).

**Table 3:**
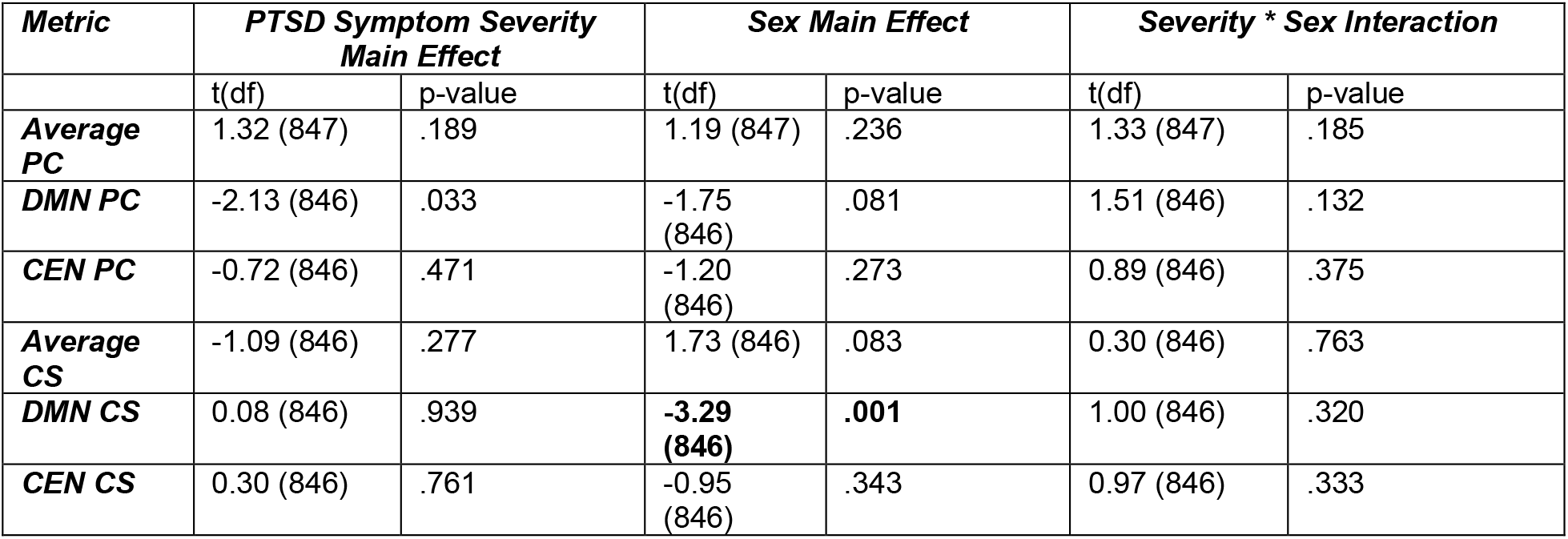
**Statistics from linear mixed effects models predicting network metrics from PTSD symptom severity and binary sex**. *PC = participation coefficient; CS = connectivity strength; DMN = default mode network; CEN = central executive network; PTSD = posttraumatic stress disorder clinical group; NTC = non-trauma exposed control group; TEC = trauma-exposed control group*. ***Bolded*** *values in table indicate significant differences after correction for multiple comparisons*.

### 3.5 Relationship between PTSD Symptom Severity, Index Trauma Type, and Graph Connectivity

LMEMs testing the moderating relationship between PTSD symptom severity and index trauma type on average whole-brain, DMN, and CEN PC and connectivity strength in the trauma-exposed groups revealed one significant main effect of index trauma type in the CEN such that individuals with IPV exposure demonstrated a reduced PC of the CEN compared to individuals in the “other” category (*t*(701)=2.52, *p*=.012; **Table 4**; **Figure 4F**).

**Table 4:**
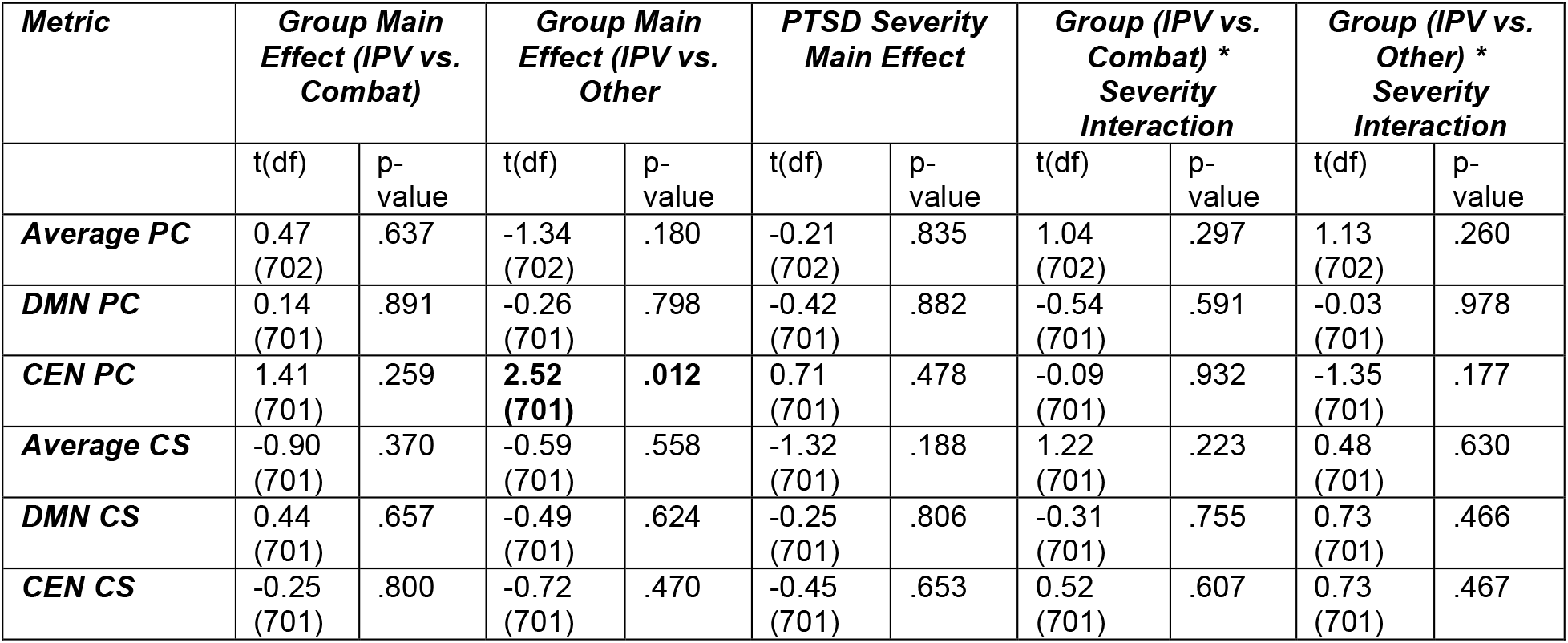
**Group-level statistics from linear mixed effects models predicting network metrics from index trauma type and PTSD symptom severity**. *PC = participation coefficient; CS = connectivity strength; DMN = default mode network; CEN = central executive network; PTSD = posttraumatic stress disorder clinical group; NTC = non-trauma exposed control group; TEC = trauma-exposed control group; IPV = interpersonal violence*. ***Bolded*** *values in table indicate significant differences after correction for multiple comparisons*.

| *Metric* | *Group Main Effect (IPV vs. Combat)* |  | *Group Main Effect (IPV vs. Other)* |  | *PTSD Severity Main Effect* |  | *Group (IPV vs. Combat) \* Severity Interaction* |  | *Group (IPV vs. Other) \* Severity Interaction* |  |
| --- | --- | --- | --- | --- | --- | --- | --- | --- | --- | --- |
|  | t(df) | p-value | t(df) | p-value | t(df) | p-value | t(df) | p-value | t(df) | p-value |
| **Average PC** | 0.47<br>(702) | .637 | -1.34<br>(702) | .180 | -0.21<br>(702) | .835 | 1.04<br>(702) | .297 | 1.13<br>(702) | .260 |
| **DMN PC** | 0.14<br>(701) | .891 | -0.26<br>(701) | .798 | -0.42<br>(701) | .882 | -0.54<br>(701) | .591 | -0.03<br>(701) | .978 |
| **CEN PC** | 1.41<br>(701) | .259 | **2.52**<br>**(701)** | **.012** | 0.71<br>(701) | .478 | -0.09<br>(701) | .932 | -1.35<br>(701) | .177 |
| **Average CS** | -0.90<br>(701) | .370 | -0.59<br>(701) | .558 | -1.32<br>(701) | .188 | 1.22<br>(701) | .223 | 0.48<br>(701) | .630 |
| **DMN CS** | 0.44<br>(701) | .657 | -0.49<br>(701) | .624 | -0.25<br>(701) | .806 | -0.31<br>(701) | .755 | 0.73<br>(701) | .466 |
| **CEN CS** | -0.25<br>(701) | .800 | -0.72<br>(701) | .470 | -0.45<br>(701) | .653 | 0.52<br>(701) | .607 | 0.73<br>(701) | .467 |

## 4. Discussion

The present study leveraged the PGC-ENIGMA PTSD Consortium resting-state dataset and a resolution limit-free community detection approach to investigate whole-brain and community-specific topological differences that distinguish individuals with PTSD from trauma-exposed and non-trauma exposed controls. Our analyses revealed significant group-level differences in PC and connectivity strength such that individuals with PTSD demonstrated higher whole-brain, CEN, and DMN average PC and connectivity strength compared to controls. We provide evidence for differential effects of index trauma exposure on CEN PC, with IPV-exposed individuals demonstrating a lower CEN PC compared to non-exposed individuals. Our robust group-level findings in PC and connectivity strength, along with the clear network structure and high value of Surprise, suggests that resolution limit-free community detection identified functional network alterations in PTSD.

Little extant literature has specifically investigated the influence of trauma exposure and PTSD on node-level or whole-brain PC. One study in combat veterans with PTSD examined node-level PC in relation to PTSD symptom severity, revealing a marginally significant positive relationship between right insula PC and re-experiencing symptoms ^59^. Further, a study in individuals with social anxiety disorder reported a larger PC in the posterior cingulate cortex and parahippocampus, regions of the DMN, in social anxiety compared to controls ^60^. These findings suggest that a more diverse distribution of edges from key nodes throughout brain networks may characterize anxious psychopathology. Though no studies to our knowledge have examined whole-brain or community-level PC in relation to PTSD, some studies have assessed modularity, a similar measure of cross-community communication that is inversely related to PC, in trauma-exposed samples. Similar to the evidence presented in this report of reduced CEN PC in IPV-exposed individuals compared to other trauma types, in adolescent girls with IPV exposure, modularity was positively correlated with emotional abuse severity ^61^. Due to the CEN’s role in emotional regulation, reduced communication from the CEN to the rest of the brain graph in IPV-exposed individuals may contribute to increased difficulty in emotional regulation that is present in individuals with sexual assault exposure compared to other trauma exposures ^62^.

Conversely, an increased PC within specific communities such as the CEN or DMN may represent a reduction in the density of connections within modules in favor of cross-module communication. Such desegregation of functional networks has been observed in PTSD and is proposed to contribute to attentional deficits ^63^. This dynamic may also represent a vulnerability factor for development of PTSD following non-IPV trauma, and may explain findings of increased integration of the amygdala with the DMN and hypercoupling of the dorsal anterior cingulate and mPFC with the CEN in PTSD patients ^64^. More research is needed to understand the clinical implications of changes in PC values in individuals with PTSD. This knowledge will assist in developing therapies aimed at remodeling networks closely linked to PTSD symptoms.

Connectivity strength has been extensively studied in network investigations of PTSD, with results consistently suggesting reduced connectivity strength throughout the brain in PTSD^60,65–69^. Our results contradict some previous findings; however, prior evidence does exist for increased resting-state connectivity of the DMN in youth with PTSD compared to non-traumatized controls ^70^ as well as in the CEN of adult earthquake survivors with PTSD compared to survivors without PTSD ^67^. Additionally, hypercoupling of mPFC regions with the CEN and increased integration of the DMN with the amygdala were observed in adults with PTSD during a threat processing task ^64^, suggesting that enhanced connectivity throughout the brain of PTSD patients may alter the functional role of emotion-processing networks in monitoring external threat. Additionally, our finding of higher DMN connectivity strength in females compared to males adds to recent evidence that regions closely coupled with the DMN shift more towards task-positive configurations throughout adulthood in males compared to females ^71^, indicating that females maintain the task-negative DMN function longer than males. This may be related to females showing a shallower slope in age-related connectivity decreases of the DMN compared to males ^37^, though future work is needed to understand relationships between binary sex and longitudinal changes in network connectivity.

### 4.1 Strengths and Limitations

While the large dataset from the PGC-ENIGMA Consortium allows for strongly powered hypothesis tests, the present study has limitations regarding heterogeneities across cohorts. Firstly, scans were acquired on different scanners with inconsistent acquisition sequences and parameters. We adjusted for these potential scanner-related differences using ComBat harmonization, which has been shown to mitigate such effects in resting-state datasets ^49^, and included a random effect of scanner for all LMEMs to address variance contributed by this factor. Additionally, generalization of our results is limited by heterogeneity in clinical assessment tools and characterization of comorbid psychiatric disorders across cohorts. Finally, information on comorbidities was not provided for all cohorts; therefore, we are unable to assess for network-level differences that may be attributed to other psychiatric concerns.

### 4.2 Conclusions

Our analysis represents the largest investigation of network-level resting-state organization in PTSD to date. The results presented provide evidence for altered network organization characterized by a greater distribution of connections between communities as well as greater connectivity strength throughout the brain graph and within emotion-regulation networks in PTSD individuals compared to controls. Future work is needed to further characterize other relevant network dynamics that may be contributing to PTSD symptoms, including hub structure, small-worldness, and node-specific connectivity profiles. Future work should also examine network organization during relevant cognitive and decision-making tasks to determine how changes in network topology facilitate behavioral deficits in clinical groups.

## Data Availability

Data from this analysis may be made available upon request from the PGC-ENGIMA PTSD consortium.

## Grant Support Acknowledgements

Research in this publication was supported by the National Institute of Mental Health and the National Institutes of Health under awards T32MH018931-31, F31MH122047 and T32GM007507.

## PGC-ENGIMA Project Funding Acknowledgements

Congressionally Directed Medical Research Program (W81XWH-08–2–0038: PI-Sponheim), the Department of Veterans Affairs, Rehabilitation R&D Program (I01RX000622: PI-Sponheim, IK2RX002922: PI-Disner); National Institute of Mental Health (R21MH112956: PI-Kaufman; K23MH090366-01: PI-Bruce; K01MH118467: PI – Lebois; MH119132 and MH097784: PI-Cisler; R01-MH111671-01 and VISN6 MIRECC: PI – Morey); VA Merit (1I01RX000389-01 and 1I01CX000748-01A1; PI-Morey) (to RAM)Grady Trauma Project supported through MH098212; MH071537; M01RR00039; UL1TR000454; HD071982; HD085850; MH101380; Narsad Young Investigator; German Research Foundation (DA 1222/4-1: PI – Daniels, WA 1539/8-2: PI – Walter); EU Rosalind-Franklin Fellowship Program (Daniels); German National Merit Foundation (Sierk). The COPTSD study was funded by a Hospital Clinical Research Program (Promoteur CHRU de Tours). We acknowledge the ‘Fondation Pierre Deniker’ and the ‘SFR FED4226 Neuroimagerie Fonctionnelle’ for their financial support. This study was also supported by a postgraduate scholarship from the French Ministry of Higher Education, Research and Innovation awarded to YQ. Canadian Institutes of Health Research (Densmoore); and Canadian Institute for Military and Veteran Health Research (Densmoore and Lanius).

## References

1. Singewald, N. & Holmes, A. Rodent models of impaired fear extinction. Psychopharmacology 236, 21–32 (2019).

2. Garcia & Vouimba, R. The amygdala modulates prefrontal cortex activity relative to conditioned fear. Nature 402, 294 (1999).

3. Morgan, M. A. & LeDoux, J. E. Differential contribution of dorsal and ventral medial prefrontal cortex to the acquisition and extinction of conditioned fear in rats. Behavioral Neuroscience 109, 681–688 (1995).

4. Pitman, R. K. et al. Biological studies of post-traumatic stress disorder. Nature Reviews Neuroscience 13, 769–787 (2012).

5. Patel, R., Spreng, R. N., Shin, L. M. & Girard, T. A. Neurocircuitry models of posttraumatic stress disorder and beyond: A meta-analysis of functional neuroimaging studies. Neuroscience & Biobehavioral Reviews 36, 2130–2142 (2012).

6. Admon, R., Milad, M. R. & Hendler, T. A causal model of post-traumatic stress disorder: disentangling predisposed from acquired neural abnormalities. Trends in Cognitive Sciences 17, 337–347 (2013).

7. Koenigs, M. & Grafman, J. Posttraumatic stress disorder: the role of medial prefrontal cortex and amygdala. Neuroscientist 15, 540–548 (2009).

8. Koenigs, M. et al. Focal brain damage protects against post-traumatic stress disorder in combat veterans. Nat Neurosci 11, 232–237 (2008).

9. Zielinski, M. J. et al. Does development moderate the effect of early life assaultive violence on resting-state networks? An exploratory study. Psychiatry Research: Neuroimaging 281, 69–77 (2018).

10. Thomason, M. E. et al. Altered amygdala connectivity in urban youth exposed to trauma. Soc Cogn Affect Neurosci 10, 1460–1468 (2015).

11. Liberzon, I. & Abelson, J. L. Context Processing and the Neurobiology of Post-Traumatic Stress Disorder. Neuron 92, 14–30 (2016).

12. Ross, M. C. & Cisler, J. M. Altered large-scale functional brain organization in posttraumatic stress disorder: A comprehensive review of univariate and network-level neurocircuitry models of PTSD. NeuroImage: Clinical 27, 102319 (2020).

13. Menon, V. Large-scale brain networks and psychopathology: a unifying triple network model. Trends in Cognitive Sciences 15, 483–506 (2011).

14. Menon, V. Developmental pathways to functional brain networks: emerging principles. Trends in Cognitive Sciences 17, 627–640 (2013).

15. Bressler, S. L. & Menon, V. Large-scale brain networks in cognition: emerging methods and principles. Trends Cogn Sci 14, 277–290 (2010).

16. Menon, V. & Uddin, L. Q. Saliency, switching, attention and control: a network model of insula function. Brain Structure and Function 214, 655–667 (2010).

17. Scolari, M., Seidl-Rathkopf, K. N. & Kastner, S. Functions of the human frontoparietal attention network: Evidence from neuroimaging. Curr Opin Behav Sci 1, 32–39 (2015).

18. Szczepanski, S. M., Konen, C. S. & Kastner, S. Mechanisms of Spatial Attention Control in Frontal and Parietal Cortex. Journal of Neuroscience 30, 148–160 (2010).

19. Sripada, C. et al. Volitional regulation of emotions produces distributed alterations in connectivity between visual, attention control, and default networks. Neuroimage 89, 110–121 (2014).

20. Wessing, I., Rehbein, M. A., Postert, C., Fürniss, T. & Junghöfer, M. The neural basis of cognitive change: Reappraisal of emotional faces modulates neural source activity in a frontoparietal attention network. NeuroImage 81, 15–25 (2013).

21. Koster, E. H. W., Crombez, G., Verschuere, B., Van Damme, S. & Wiersema, J. R. Components of attentional bias to threat in high trait anxiety: Facilitated engagement, impaired disengagement, and attentional avoidance. Behaviour Research and Therapy 44, 1757–1771 (2006).

22. Sippel, L. M. & Marshall, A. D. Posttraumatic Stress Disorder and Fear of Emotions: The Role of Attentional Control. J Trauma Stress 26, 397–400 (2013).

23. Raichle, M. E. The Brain’s Default Mode Network. Annual Review of Neuroscience 38, 433–447 (2015).

24. Sporns, O. & Betzel, R. F. Modular Brain Networks. Annu Rev Psychol 67, 613–640 (2016).

25. Guimerà, R. & Nunes Amaral, L. A. Functional cartography of complex metabolic networks. Nature 433, 895–900 (2005).

26. Bassett, D. S. & Sporns, O. Network neuroscience. Nature Neuroscience 20, 353–364 (2017).

27. Fortunato, S. Community detection in graphs. Physics Reports 486, 75–174 (2010).

28. Fortunato, S. & Barthelemy, M. Resolution limit in community detection. Proceedings of the National Academy of Sciences 104, 36–41 (2007).

29. Newman, M. E. J. & Girvan, M. Finding and evaluating community structure in networks. Physical Review E 69, (2004).

30. Pedersen, M., Omidvarnia, A., Shine, J. M., Jackson, G. D. & Zalesky, A. Reducing the influence of intramodular connectivity in participation coefficient. Netw Neurosci 4, 416–431 (2020).

31. Bordier, C., Nicolini, C., Forcellini, G. & Bifone, A. Disrupted modular organization of primary sensory brain areas in schizophrenia. Neuroimage Clin 18, 682–693 (2018).

32. Kessler, R. C. et al. Trauma and PTSD in the WHO World Mental Health Surveys. Eur J Psychotraumatol 8, 1353383 (2017).

33. Kilpatrick, D. G. et al. National Estimates of Exposure to Traumatic Events and PTSD Prevalence Using DSM-IV and DSM-5 Criteria: DSM-5 PTSD Prevalence. Journal of Traumatic Stress 26, 537– 547 (2013).

34. Breslau, N., Davis, G. C., Andreski, P., Peterson, E. L. & Schultz, L. R. Sex differences in posttraumatic stress disorder. Arch. Gen. Psychiatry 54, 1044–1048 (1997).

35. Olff, M. Sex and gender differences in post-traumatic stress disorder: an update. European Journal of Psychotraumatology 8, 1351204 (2017).

36. Tian, L., Wang, J., Yan, C. & He, Y. Hemisphere-and gender-related differences in small-world brain networks: A resting-state functional MRI study. NeuroImage 54, 191–202 (2011).

37. Scheinost, D. et al. Sex differences in normal age trajectories of functional brain networks. Human Brain Mapping 36, 1524–1535 (2015).

38. Blake, D. D. et al. The development of a Clinician-Administered PTSD Scale. J Trauma Stress 8, 75– 90 (1995).

39. Weathers, F. W. et al. The Clinician-Administered PTSD Scale for DSM–5 (CAPS-5): Development and initial psychometric evaluation in military veterans. Psychological Assessment 30, 383–395 (2018).

40. Blanchard, E. B., Jones-Alexander, J., Buckley, T. C. & Forneris, C. A. Psychometric properties of the PTSD checklist (PCL). Behaviour Research and Therapy 34, 669–673 (1996).

41. Blevins, C. A., Weathers, F. W., Davis, M. T., Witte, T. K. & Domino, J. L. The Posttraumatic Stress Disorder Checklist for DSM-5 (PCL-5): Development and Initial Psychometric Evaluation. J Trauma Stress 28, 489–498 (2015).

42. Foa, E. B., Cashman, L., Jaycox, L. & Perry, K. The validation of a self-report measure of posttraumatic stress disorder: The Posttraumatic Diagnostic Scale. Psychological Assessment 9, 445–451 (1997).

43. Waller, L. et al. ENIGMA HALFpipe: Interactive, reproducible, and efficient analysis for resting-state and task-based fMRI data. bioRxiv 2021.05.07.442790 (2021) doi:10.1101/2021.05.07.442790.

44. Esteban, O. et al. fMRIPrep: a robust preprocessing pipeline for functional MRI. Nature methods 16, (2019).

45. Pruim, R. H. R. et al. ICA-AROMA: A robust ICA-based strategy for removing motion artifacts from fMRI data. NeuroImage 112, 267–277 (2015).

46. Cox, R. W. AFNI: software for analysis and visualization of functional magnetic resonance neuroimages. Comput. Biomed. Res. 29, 162–173 (1996).

47. Craddock, R. C., James, G. A., Holtzheimer, P. E., Hu, X. P. & Mayberg, H. S. A whole brain fMRI atlas generated via spatially constrained spectral clustering. Hum Brain Mapp 33, 1914–1928 (2012).

48. Fortin, J.-P. et al. Harmonization of cortical thickness measurements across scanners and sites. NeuroImage 167, 104–120 (2018).

49. Yu, M. et al. Statistical harmonization corrects site effects in functional connectivity measurements from multi-site fMRI data. Hum Brain Mapp 39, 4213–4227 (2018).

50. Nicolini, C., Bordier, C. & Bifone, A. Community detection in weighted brain connectivity networks beyond the resolution limit. NeuroImage 146, 28–39 (2017).

51. Gallos, L. K., Makse, H. A. & Sigman, M. A small world of weak ties provides optimal global integration of self-similar modules in functional brain networks. Proc Natl Acad Sci U S A 109, 2825– 2830 (2012).

52. Aldecoa, R. & Marín, I. Deciphering Network Community Structure by Surprise. PLOS ONE 6, e24195 (2011).

53. Aldecoa, R. & Marín, I. Surprise maximization reveals the community structure of complex networks. Sci Rep 3, 1060 (2013).

54. Nicolini, C. & Bifone, A. Modular structure of brain functional networks: breaking the resolution limit by Surprise. Scientific Reports 6, 19250 (2016).

55. Jaccard, P. Étude comparative de la distribution florale dans une portion des Alpes et des Jura. Bulletin de la Société vaudoise des sciences naturelles 37, 547–579 (1901).

56. Yeo, B. T. et al. The organization of the human cerebral cortex estimated by intrinsic functional connectivity. J Neurophysiol 106, 1125–1165 (2011).

57. Rubinov, M. & Sporns, O. Complex network measures of brain connectivity: Uses and interpretations. NeuroImage 52, 1059–1069 (2010).

58. Holm, S. A Simple Sequentially Rejective Multiple Test Procedure. Scandinavian Journal of Statistics 6, 65–70 (1979).

59. Spielberg, J. M., McGlinchey, R. E., Milberg, W. P. & Salat, D. H. Brain network disturbance related to posttraumatic stress and traumatic brain injury in veterans. Biol. Psychiatry 78, 210–216 (2015).

60. Yang, X. et al. Network analysis reveals disrupted functional brain circuitry in drug-naive social anxiety disorder. NeuroImage 190, 213–223 (2019).

61. Cisler, J. M. Childhood Trauma and Functional Connectivity between Amygdala and Medial Prefrontal Cortex: A Dynamic Functional Connectivity and Large-Scale Network Perspective. Front. Syst. Neurosci. 11, (2017).

62. Raudales, A. M., Short, N. A. & Schmidt, N. B. Emotion dysregulation mediates the relationship between trauma type and PTSD symptoms in a diverse trauma-exposed clinical sample. Personality and Individual Differences 139, 28–33 (2019).

63. Block, S. R. & Liberzon, I. Attentional processes in posttraumatic stress disorder and the associated changes in neural functioning. Exp Neurol 284, 153–167 (2016).

64. Rabellino, D. et al. Intrinsic Connectivity Networks in post-traumatic stress disorder during sub- and supraliminal processing of threat-related stimuli. Acta Psychiatr Scand 132, 365–378 (2015).

65. Akiki, T. J. et al. Default mode network abnormalities in posttraumatic stress disorder: A novel network-restricted topology approach. Neuroimage 176, 489–498 (2018).

66. Puetz, V. B. et al. Altered brain network integrity after childhood maltreatment: A structural connectomic DTI-study. Human Brain Mapping 38, 855–868 (2017).

67. Shang, J. et al. Alterations in Low-Level Perceptual Networks Related to Clinical Severity in PTSD after an Earthquake: A Resting-State fMRI Study. PLoS One 9, (2014).

68. Tursich, M. et al. Distinct intrinsic network connectivity patterns of post-traumatic stress disorder symptom clusters. Acta Psychiatrica Scandinavica 132, 29–38 (2015).

69. Roos, A., Fouche, J.-P. & Stein, D. J. Brain network connectivity in women exposed to intimate partner violence: a graph theory analysis study. Brain Imaging and Behavior 11, 1629–1639 (2017).

70. Patriat, R., Birn, R. M., Keding, T. J. & Herringa, R. J. Default-Mode Network Abnormalities in Pediatric Posttraumatic Stress Disorder. Journal of the American Academy of Child & Adolescent Psychiatry 55, 319–327 (2016).

71. Conrin, S. D. et al. From Default Mode Network to the Basal Configuration: Sex Differences in the Resting-State Brain Connectivity as a Function of Age and Their Clinical Correlates. Front. Psychiatry 9, (2018).

