## Supplementary material for "Resolution limit-free community detection reveals unique patterns of resting-state network connectivity in posttraumatic stress disorder: A PGC-ENIGMA PTSD Consortium investigation": Tables S1 and S2

**Table S1**: Summary of PTSD Diagnosis and Demographics of Individual Cohorts.

| | | | ***PTSD Diagnosis*** | | | | ***Binary Sex*** | | ***Age*** | |
|---|---|---|---|---|---|---|---|---|---|---|
| **Site** | **Cohort/Scanner** | **N** | **PTSD** | **TEC** | **NTC** | **Instrument** | **Female** | **Male** | **Mean** | **SD** |
| AMC | BOOSTER | 58 | 28 | 30 | 0 | CAPS-IV | 31 | 27 | 36.2 | 7.3 |
| UAMS | DOP | 26 | 26 | 0 | 0 | CAPS-5 | 26 | 0 | 38.0 | 7.2 |
| UAMS | PAL | 25 | 10 | 0 | 15 | PCL-C | 27 | 0 | 31.4 | 7.0 |
| Wisconsin | DOP | 43 | 43 | 0 | 0 | CAPS-5 | 43 | 0 | 30.9 | 8.6 |
| Wisconsin | EMOREG | 12 | 1 | 0 | 11 | CAPS-5 | 12 | 0 | 32.7 | 8.3 |
| Columbia* | MR750 | 48 | 18 | 15 | 15 | CAPS-5 | 21 | 27 | 30.7 | 9.5 |
| Columbia | Premier | 12 | 0 | 2 | 10 | CAPS-5 | 8 | 4 | 30.2 | 7.9 |
| Emory | GTP | 85 | 27 | 57 | 1 | DSM-IV | 85 | 0 | 34.2 | 8.4 |
| Gronigen | DISPO | 32 | 32 | 0 | 0 | CAPS-IV | 32 | 0 | 36.3 | 8.6 |
| Masaryk | Holocaust Survivors | 63 | 0 | 0 | 63 | PCL-C | 35 | 28 | 33.8 | 8.6 |
| McLean | | 64 | 41 | 20 | 3 | CAPS-5 | 64 | 0 | 30.4 | 8.9 |
| Michigan | Mindfulness | 56 | 38 | 0 | 18 | CAPS-IV | 0 | 56 | 31.3 | 7.8 |
| Milwaukee | Larson | 78 | 17 | 61 | 0 | CAPS-5 | 45 | 33 | 31.8 | 9.1 |
| Minneapolis VA | SATURN | 69 | 35 | 34 | 0 | CAPS-IV | 5 | 64 | 32.7 | 7.6 |
| UK Muenster | Munster | 44 | 17 | 0 | 27 | PDS | 35 | 9 | 25.3 | 5.9 |
| Stanford | Brains | 71 | 45 | 24 | 0 | CAPS-IV | 40 | 31 | 33.6 | 8.4 |
| Stanford | CausCon | 60 | 0 | 56 | 4 | CAPS-IV | 43 | 17 | 30.8 | 9.4 |
| Tours | COPTSD | 38 | 8 | 11 | 19 | CAPS-IV | 30 | 0 | 26.3 | 8.1 |
| Toledo* | MVA | 39 | 5 | 34 | 0 | CAPS-IV | 29 | 10 | 30.3 | 8.8 |
| Toledo* | ONG | 27 | 6 | 21 | 0 | CAPS-IV | 24 | 3 | 36.7 | 9.0 |
| Utrecht* | BETER | 57 | 30 | 15 | 12 | CAPS-IV | 0 | 57 | 33.6 | 8.5 |
| Wisconsin | Veteran’s Wellness | 58 | 19 | 39 | 0 | CAPS-IV | 4 | 54 | 30.7 | 6.6 |
| Waco Veteran’s Hospital | MAVRIX | 28 | 14 | 14 | 0 | PCL-5 | 3 | 25 | 36.4 | 7.5 |
| Waco Veteran’s Hospital | TEMI | 10 | 10 | 0 | 0 | PCL-5 | 0 | 10 | 37.1 | 5.7 |
| Western Ontario | Lawson | 131 | 0 | 45 | 86 | CAPS-IV | 87 | 44 | 33.5 | 10.2 |

* Cohort not included in main analyses

**Table S2**: Summary of Inclusion and Exclusion Criteria for Individual Cohorts

| Site | Cohort | Inclusion Criteria | Exclusion Criteria |
|---|---|---|---|
| AMC | BOOSTER | PTSD patients had to fulfill the DSM-IV diagnostic criteria for PTSD, with a score of > 45 on the clinician-administered PTSD scale (CAPS). | PTSD patients were excluded if they met DSM-IV criteria for current psychotic disorder, substance-related disorder, severe personality disorder, severe major depressive disorder (MDD) (ie, involving high suicidal risk and/or psychotic symptoms) or current suicidal risk |
| UAMS | DOP | Age 21-50, fluent in English, experience of interpersonal violence, meets criteria for PTSD diagnosis on CAPS | Psychotic symptoms, past psychotic disorders, stable on medications < 4 weeks, cognitive impairment, current substance or alcohol use disorder, MRI contraindications |
| UAMS | PAL | Age 21-50, fluent in English, experience of interpersonal violence (for PTSD group) | Psychotic symptoms, past psychotic disorders, stable on medications < 4 weeks, cognitive impairment, current substance or alcohol use disorder, MRI contraindications |
| Wisconsin | DOP | Age 21-50, fluent in English, experience of interpersonal violence, meets criteria for PTSD diagnosis on CAPS | Psychotic symptoms, past psychotic disorders, stable on medications < 4 weeks, cognitive impairment, current substance or alcohol use disorder, MRI contraindications |
| Wisconsin | EMOREG | Age 21-50, fluent in English, experience of interpersonal violence (for PTSD group) | Psychotic symptoms, past psychotic disorders, stable on medications < 4 weeks, cognitive impairment, current substance or alcohol use disorder, MRI contraindications |
| Columbia* | MR750 | Between the ages of 18 and 60. Experience of a traumatic event or events in childhood and/or adulthood; current DSM-V Criterion A for PTSD. Able to give consent, fluent in English | Prior or current Axis I psychiatric diagnosis of schizophrenia, psychotic disorder, bipolar disorder, dementia. Depression score of > 25 on the Hamilton Rating Scale for Depression (HAM-D-17-item); significant depression and /or depression related impairment that is judged to warrant pharmacotherapy or combined medication and psychotherapy.Individuals at risk for suicide based on history and current mental state. History of substance/alcohol dependence within the past six months, or abuse within past two months. Any psychotropic medications. Pregnancy, or plans to become pregnant during the period of the study. Paramagnetic metallic implants or devices contraindicating magnetic resonance imaging or any other non-removable paramagnetic metal in the body. Medical illness that could interfere with assessment of diagnosis, or biological measures (SCR, fMRI), including organic brain impairment from stroke, CNS tumor, or demyelinating disease; and renal, thyroid, hematologic or hepatic |

| | | | |
|---|---|---|---|
| | | | impairment. Any condition that would exclude MRI exam (e.g. pacemaker, paramagnetic metallic prosthesis, surgical clips, shrapnel, necessity for constant medicinal patch, some tattoos) |
| Columbia | Primier | Between the ages of 18 and 60. Experience of a traumatic event or events in childhood<br>and/or adulthood; current DSM-V Criterion A for PTSD. Able to give consent, fluent in English | Prior or current Axis I psychiatric diagnosis of schizophrenia, psychotic disorder, bipolar disorder, dementia. Depression score of > 25 on the Hamilton Rating Scale for Depression (HAM-D-17-item); significant depression and /or depression related impairment that is judged to warrant pharmacotherapy or combined medication and psychotherapy.Individuals at risk for suicide based on history and current mental state. History of substance/alcohol dependence within the past six months, or abuse within past two months. Any psychotropic medications. Pregnancy, or plans to become pregnant during the period of the study. Paramagnetic metallic implants or devices contraindicating magnetic resonance imaging or any other non-removable paramagnetic metal in the body.Medical illness that could interfere with assessment of diagnosis, or biological measures (SCR, fMRI), including organic brain impairment from stroke, CNS tumor, or demyelinating disease;<br>and renal, thyroid, hematologic or hepatic impairment. Any condition that would exclude MRI exam (e.g. pacemaker, paramagnetic metallic prosthesis, surgical clips, shrapnel, necessity for constant medicinal patch, some tattoos) |
| Emory | GTP | 18-65 years of age, endorsed at least 1 criterion A trauma, English-speaking | Current psychotic symptoms or bipolar disorder, current substance or alcohol dependence, history of head trauma, taking any psychoactive medication, current illegal drug use (verified with urine drug screen within 24 hours of scan) |
| Gronigen | DISPO | Age 20-60, ALL FEMALES with trauma exposure before age 21; Diagnostic interviews by Clinician with CAPS, SCID-1, SCID-2, and SCID-D; all subjects have PTSD diagnosis (according to CAPS), fluent in German, | Psychotic disorders, dissociative identity disorder, current alcohol dependency, current substance intoxication, ferrous metal in the body, neurological disorders, history of TBI, claustrophobia |
| Masaryk | Holocaust Survivors | Age 15-95, fluent in Czech/Slovak & capable of understanding consent, score in MMSE over 26 | Neurological disorders, claustrophobia, Psychotic disorders, ferrous metal in the body |
| McLean | Traumatic Dissociation | Patients: Female; Diagnosis of PTSD; history of childhood abuse; Age between 18 and 89; Legal and mental competency of the patient | Male;<br>Under 18 or over 89;<br>Legal or mental incompetence;<br>Delirium secondary to medical illness;<br>PTSD or DID due to general medical or neurological illness; |

| | | | |
|---|---|---|---|
| | | Controls: Female; no history of psychiatric illness; Age between 18 and 89 | History of neurological conditions that may cause significant psychiatric symptomatology (e.g., dementia;)<br>Any contraindication to MR scans, including claustrophobia, pregnancy, metal implants, etc.;<br>Current alcohol or substance dependence or abuse (within the last month);<br>A history of schizophrenia or other psychotic disorder;<br>History of head injury or loss of consciousness for longer than 5 min (including concussion) |
| Michigan | Mindfulness | Age 18-65, fluent in English & capable of understanding consent, OEF/OIF Veteran | Axis I disorders (except Depression, GAD, PTSD, Panic Disorder, Agoraphobia, Other Specific Phobias, Anxiety NOS), neurological disorders, Current or history of Psychotic disorders, Suicide attempts in past year, ferrous metal in the body,, claustrophobia, or other contraindication for MRI |
| Milwaukee | Larson | PTSD criterion A met, age 18-60, GCS>= 13 (mild TBI criteria), Rothbaum 3 or higher or item 2 rated 3 or higher, English speaking (either native or bilingual proficiency), able to schedule within 30 days of brain injury. | Still in high school, re-admitted to hospital for current brain injury, live too far away to travel for study, police hold, incarcerated, intentional self-inflicted injury, known perpetrator, moderate to severe cognitive impairment, loss of consciousness > 30 minutes, pregnant, clear evidence of substance abuse, anti-psychotic or anti-seizure medication, indication of psychotic disorder or manic symptoms, MRI contraindications, history of seizures or other neurological conditions, severe hearing or vision problems |
| Minneapolis VA | SATURN | age: 18-60, OEF/OIF, deployed | Moderate/severe TBI, non-TBI neurological conditions, current psychotic symptoms, substance abuse/dependence other than alcohol, unstable med conditions, sig risk of suicide/homicide |
| UK Muenster | Munster | All patients fulfilled the diagnostic criteria for PTSD as primary diagnosis according to the DSM-IV-TR (American Psychiatric Association, 2000), assessed by the German version of the Structured Clinical Interview for DSM-IV (SCID; Wittchen et al., 1997).<br>Given the focus on IPV-PTSD, the experience of a trauma related to IPV (e.g., rape, sexual or physical abuse) at least once was an inclusion criterion for the patient group. All participants had normal or corrected-to-normal vision and were right-handed as determined by the Edinburgh Handedness Inventory (Oldfield, 1971).<br>Written informed consent was obtained from all participants, who were recruited via public announcements in newspapers | No control had a lifetime PTSD, MRI contraindications |

| | | | |
|---|---|---|---|
| | | and received reimbursement for their participation | |
| Stanford | BRAINS | Age 18-65, fluent in English & capable of understanding consent, OEF/OIF Veteran | History of psychotic, bipolar or substance dependence (within 3 months for patients and lifetime for controls), a history of a neurological disorder, greater than mild traumatic brain injury (i.e. >30 minutes loss of consciousness or >24 hour post-trauma amnesia), claustrophobia, and regular use of benzodiazepines, opiates, thyroid medications, or other CNS medication. Trauma-exposed healthy controls were required to have experienced a criterion A trauma, but not meet lifetime criteria for any Axis 1 psychiatric disorder, including PTSD. |
| Stanford | CausCon | Patients will be required to have chronic(>3 months)moderate to severe anxiety or depression, assessed dimensionally by a score on the PHQ9 scale(excluding the suicide question)>10 or a score on the GAD7scale >10. Both of these scales assess general symptoms of anxiety and depression, and these cutoffs have been shown to relate to moderate or greater severity of symptoms. Moreover, because these scales measure general anxiety and depression, they are sensitive to a wide range of DSM diagnoses, including GAD, MD and PTSD. Additionally, to ensure clinical significance, subjects will need to indicate that they would be interested in seeking treatment for these symptoms(i.e. that symptoms impair functioning). Other inclusion criteria are:(1)community dwelling adults ages 18-60 years old;(2)not currently in treatment;(3)free of metal or ferrous implant,(4)good English comprehension and non-impaired intellectual abilities to ensure understanding of task instructions;(5)no history of neurological disorders, brain surgery, electroconvulsive or radiation treatment, brain hemorrhage or tumor, stroke, epilepsy, hypo- or hyperthyroidism, and(6) no daily use of PRN benzodiazepines or opiates(max: 3x/wk), or daily thyroid medications, and no antidepressant, anticonvulsant or antipsychotic medications for > 2 wks(fluoxetine >6 wks). As-needed benzodiazepines or opiates cannot be used within 48 hours of assessments. Medication-free healthy subjects will likewise be split equally between those who have never been traumatized and those who have had a criterion A trauma. Controls must deny lifetime psychiatric | 1. MRI counter-indications(e.g. shrapnel or other metal in/on the body that can not be removed, claustrophobia, etc.)2. Additional TMS counter-indications(seizure disorder, CNS active disorder, certain medications described below)3. Medication use that substantially reduces seizure threshold to TMS(olanzapine, chlorpromazine, lithium)and unwilling or unable medically(determined by patient and his/her physician)to safely withdraw, at least two weeks prior to TMS, from these medications4. Opiate medication, antihypertensive medication, or any medication that interferes with blood flow(interferes with fMRI recordings)5. Thyroid dysfunction not adequately controlled by medication6. History of neurological or cardiovascular disorders, brain surgery, radiation treatment, brain hemorrhage or tumor, stroke, or diabetes7. Diagnosis of substance dependence within the past 3 months(but not abuse)8. Refusal to abstain from illicit drug use for duration of the study9. Refusal to abstain from alcohol within 24 hours of scans10. Pregnancy in female participants11. Prior exposure to deep brain stimulation, rTMS, or tDCS(transcranial direct current stimulation)therapies12. Significant traumatic brain injury(loss of consciousness, post-injury amnesia, significant radiological/neurological findings, penetrating brain injury)13. Lifetime evidence of psychosis, mania, hypomania, or bipolar disorders on the SCID. |

| | | | |
|---|---|---|---|
| | | diagnosis and treatment and have PHQ9 andGAD7≤4. Stratification of each group by trauma exposure will be re-assessed every 20 participants and wewill ensure that groups are matched on demographic variables. | |
| Tours | COPTSD | Age 18-65, fluent in French, scanning time at 6 months post exposure to sexual assault. | History of head injury, substance use, claustrophobia, current use of a psychotropic medication for more than 21 days, medical disorders affecting brain function (e.g., epi- lepsy, tumour), and MRI contraindications. |
| Toledo* | MVA | Survivors of a Motor Vehicle Accident (MVA) who are transported to the University of Toledo Emergency department, or to a ProMedica emergency medicine department. | Pregnancy; under the influence of alcohol or drugs at the time of MVA; major injuries, moderate to severe traumatic brain injury; major medical illnesses; conditions affecting ability to undergo MRI scans. |
| Toledo* | ONG | Ohio National Guard and Reserve soldiers, deployment in OEF or OIF; 18-50 years of age, meeting Ohio National Guard Study characteristics, and able to give informed consent. | a) life history of a neurologic condition, psychosis or bipolar disorder, b) active substance dependence, c) organic mental syndrome or pervasive developmental disorder; d) presence of ferrous metal in the body (e.g., aneurysm clips, shrapnel); or e) no psychotropic medication is preferred while use of antipsychotics will be excluded. |
| Utrecht* | BETER | All: 18-60 years of age, eligible for MRI. PTSD: current PTSD diagnosis, with CAPS ≥ 45, military deployment >4 months. Trauma controls: exposure to at least one traumatic event (according to DSM-IV A1 criterion), with CAPS < 15, no current psychiatric disorder, military deployment >4 months; healthy controls: no current psychiatric disorder according to DSM-IV. | Alcohol / drug abuse or dependency during treatment, neurological disorders (i.e. Parkinson's Disease), claustrophobia, pacemakers or other metals that might interfere with an MRI scan. |
| Wisconsin | Veteran's Wellness | Age range of 18-50.<br>Capable of giving informed consent<br>Fluent in English<br>Exposure to one or more life-threatening war zone trauma events per the Combat Experiences Scale and documented by DD-214, Combat Action Ribbon (Marines), Combat Infantry Badge (Army), or other clear evidence of war zone trauma exposure in Iraq or Afghanistan since 2001<br>Pharmacological or psychotherapeutic treatment stable for at least 8 weeks prior to beginning of study, with no intent to begin a new course of treatment during the study period | Medical<br>Weight of 352 pounds or over (due to constraints of MRI scanner)<br>Women of childbearing potential with positive pregnancy test, looking to conceive during the research timeline, or who are breastfeeding<br>Metallic implants such as prostheses or aneurysm clip, or electronic implants such as cardiac pacemakers<br>Neurological or serious medical condition that may contraindicate MRI or that may overlap with physiological substrates of psychiatric conditions<br>History of seizures or seizure disorder<br>Moderate or severe traumatic brain injury (over 20 minutes unconscious)<br><br>Psychiatric/Behavioral<br>Current active substance dependence or dependence within 3 months (other than |

| | | | |
|---|---|---|---|
| | | | nicotine)<br>Meets DSM-IV criteria for bipolar disorder, schizophrenia, schizoaffective disorder, psychotic disorder NOS, delirium, or any DSM-IV cognitive disorder.<br>Substance dependence disorder within 3 months or any current substance dependence<br>Severe psychiatric instability or severe situational life crises, including evidence of being actively suicidal or homicidal, or any behavior that poses an immediate danger to patient or others.<br>Participants with extensive experience in yoga or meditation<br><br>Medications/Therapies<br>Current use of benzodiazepines and beta-blockers |
| Waco Veteran's Hospital | MAVRIX | Veteran, age 18-60, agreement to donate saliva | Serious general medical condition that would risk the subject being able to complete MRI (active seizure disorder, dementia, active back or muscle spasms), MRI safety screen positive (metal) or history of penetrating head or eye wound without subsequent radiological evidence that the wound is metal-free. Subjects that are (were) welders or subjects that have had metal surgically removed from their eyes will not be allowed to participate without subsequent radiological evidence that the wound is metal-free, MRI quality problems (tremors, significant claustrophobia, teeth braces) |
| Waco Veteran's Hospital | TEMI | Male and female Veterans enrolled in a CTVHCS PTSD treatment program who are 18-60 years old. | (1) pregnancy; (2) exposure to metal in the eyes; (3) shrapnel or other metal embedded in the body; (4) ferromagnetic surgical implants; (5) mechanical implants (e.g., pacemakers); (6) electrical implants (e.g., cochlear implants); (7) non-removable metallic devices (e.g. stables, neck braces, or artificial limbs) (8) tattoos not done professionally; (9) non-removable body piercings; (10) current psychosis including Axis I psychotic disorder, bipolar disorder, or schizophrenia; (11) dementia or another severe cognitive disorder; (12) prior exposure to an rTMS or dTMS; (13) seizure disorder; (14) positive screen for suicidal intent, plan, or behavior within the past 6 months; (15) a TMS motor threshold of 70% or greater of the machine's maximum output. |
| Western Ontario | Lawson | Primary diagnosis of PTSD for patients | Incompatibilities with scanning conditions, previous neurologic and development illness, comorbid schizophrenia or bipolar disorder, alcohol or substance abuse, a history of head trauma, or pregnancy |

| | | | |
|---|---|---|---|
| | | | during scan. participants were excluded if they had implants or metal that do not comply with 3T fMRI safety standards for research, a history of head injury with a loss of consciousness, significant untreated medical illness, a history of neurological disorders, history of any pervasive developmental disorders, pregnancy, and current use of any psychotropic medication within one month prior to study. PTSD individuals were further excluded if they reported a history of bipolar disorder, schizophrenia, or substance-use disorder prior to participation of the study |

* Cohort not included in main analyses

**Table S3**: Resting State Scan Acquisition Information for Individual Cohorts.

| Cohort | Scanner Type | Scanner Model | Coil Channels | Voxel Size (mm) | FOV (mm) | AqOr | TR (s) | TE (ms) | Flip Angle | Number of Slices | Scan Time (min) | Eyes Open or Closed |
|---|---|---|---|---|---|---|---|---|---|---|---|---|
| AMC BOOSTER | Philips | Achieva 3T | 32 | 3 x 3 x 3 | 240x240 | Axial | 2 | 28 | 76 | 37 | 8 | Open |
| Cisler UAMS DOP and PAL | Philips | Achieva 3T X-Series | 32 | 3 x 3 x 3 | 240x240 | Oblique | 2 | 30 | 90 | 37 | 7.5 | Open |
| Cisler UW DOP and EMOREG | GE | MR750 3T | 8 | 4 x 3.75 x 3.75 | 240x240 | Sagittal | 2 | 25 | 60 | 40 | 7.5 | Open |
| Columbia | GE | MR750 3T | 32 | 3 x 3 x 4 | 192x192 | Interleaved | 1.3 | 28 | 60 | 27 | 6 | Open |
| Columbia | GE | Premier 3T | 32 | 3 x 3 x 4 | 192x192 | Interleaved | 1.3 | 28 | 60 | 27 | 6 | Open |
| Emory GTP | Siemens | Trio 3T | 12 | 3.4 x 3.4 x 4 | 220x220 | Interleaved | 2.95 | 30 | 90 | 30 | 7.38 | Open |
| Gronigen DISPO | Siemens | Magnetom Trio Tim 3T | 12 | 3 x 3 x 3 | 192x192 | Transverse | 2.25 | 25 | 80 | 40 | 5 | Open |
| Masaryk Holocaust Survivors | Siemens | Prisma 3T | 64 | 3 x 3 x 3 | 192x192 | NP$^{+}$ | 2.51 | 35 | 70 | 44 | 8 | Closed |
| McLean | Siemens | Trio Tim 3T | 12 | 3 x 3 x 3 | 216x216 | Transverse | 3 | 30 | 85 | 47 | 6:12 | Open |
| Michigan Mindfulness | Philips | Achieva 3T X-Series | 8 | 3 x 3 x 3 | 220x220 | Axial | 2 | 25 | 90 | 42 | 8 | Open |
| Milwaukee Larson | GE | MR750 3T | 32 | 3.5 x 3.5 x 3.5 | 224x224 | Sagittal | 2 | 25 | 77 | 41 | 8 | Open |
| Minneapolis VA SATURN | Siemens | Trio 3T | 12 | 3.4 x 3.4 x 4 | 220x220 | Axial | 2 | 30 | 90 | 34 | 6 | Closed |
| UK Muenster | Siemens | Magnetom Prisma 3T | 20 | 2.3 x 2.3 x 3 | 208x208 | Axial | 2.08 | 30 | 90 | 36 | 7 | Closed |

| | | | | | | | | | | | | |
|---|---|---|---|---|---|---|---|---|---|---|---|---|
| Stanford BRAINS and CausCon | GE | MR750 3T | 8 | 3.4 x 3.4 x 4.9 | 220x220 | Axial | 2 | 30 | 80 | 29 | 8 | Open |
| Tours COPTSD | Siemens | Magnetom Verio 3T | 12 | 3.3 x 3.3 x 3.3 | 60x64 | Transverse | 3 | 30 | 80 | 45 | 6:38 | Closed |
| Toledo MVA* | GE | Signa HDxt 3T | NP+ | 3.75 x 3.75 x 3.5 | 240 x 240 | Axial Interleaved | 2 | 30 | 90 | 34 | 8 | Open |
| Toledo ONG* | GE | Signa HDxt 3T | NP+ | 3.75 x 3.75 x 3.5 | 240 x 240 | Axial Interleaved | 2 | 30 | 90 | 34 | 8 | Open |
| Utrecht BETER* | Philips | Achieva 3T | 8 | 4 x 4 x 3.6 | 208 x 120 x 256 | Transverse | 1.6 | 23 | NP* | 30 | 8.75 | Open |
| Wisconsin Veteran's Wellness | GE | X750 Discovery 3T | 8 | 1.7 x 1.7 x 4 | 220x220 | Sagittal | 2 | 20 | 60 | 40 | 7 | Closed |
| Waco MAVRIX and TEMI | Philips | Achieva | 8 | 4 x 4 x 4 | 256x256 | Axial | 2.2 | 32 | 90 | 34 | 4.8 | Open |
| Lawson/ Ontario | Seimens | Biograph mMR 3T | 32 | 2 x 2 x 2 | 192x192 | Axial | 3 | 20 | 90 | NP+ | 6 | Closed |

+ NP – parameter was not provided by the acquisition site
* Cohort not included in main analyses

**Table S4:** Jackknife Analysis for Quality Control. Cohorts were excluded from final analyses if *BOTH* the ROI coverage (N Good ROIs) *and* maximum surprise value increased when they were removed during the jackknife analysis.

| **Removed Site Number** | **Sub N** | **N Good ROIs** | **Max Surprise** | **N Communities** |
|---|---|---|---|---|
| All | 1233 | 192 | 1360.8 | 20 |
| 27 | 1195 | 192 | 1349.0 | 20 |
| **26** | 1168 | ***194*** | ***1410.6*** | 20 |
| 24 | 1221 | 192 | 1351.9 | 19 |
| **23** | 1185 | ***202*** | ***1449.4*** | 21 |
| 22 | 1170 | 192 | 1340.6 | 20 |
| 21 | 1102 | 192 | 1428.3 | 19 |
| 20 | 1102 | 192 | 1346.3 | 21 |
| 19 | 1164 | 192 | 1337.2 | 20 |
| **17** | 1176 | ***200*** | ***1523.5*** | 19 |
| 16 | 1155 | 192 | 1401.4 | 19 |
| 15 | 1175 | 192 | 1328.7 | 21 |
| 14 | 1175 | 192 | 1321.7 | 21 |
| 10 | 1189 | 192 | 1335.8 | 21 |
| 9 | 1177 | 192 | 1314.4 | 20 |
| 7 | 1201 | 192 | 1359.3 | 19 |
| 6 | 1178 | 192 | 1362.2 | 21 |
| 5 | 1182 | 193 | 1336.4 | 19 |
| 3 | 1169 | 192 | 1361.7 | 18 |
| 2 | 1148 | 192 | 1334.2 | 18 |
| 1 | 1195 | 192 | 1329.9 | 22 |
